# Longitudinal immune profiling in sepsis reveals transient expansion of a CD14⁺ monocyte state and time-dependent host-response endotypes

**DOI:** 10.1101/2025.05.17.25327533

**Authors:** Pierre O. Ankomah, Alyssa DuBois, Abraham Sonny, Miguel Reyes, Aidan C. Greenaway, Alyson L. Yee, Paul C. Blainey, Marcia B. Goldberg, Michael R. Filbin, Nir Hacohen, Roby P. Bhattacharyya

## Abstract

Sepsis is a leading cause of global mortality, yet how the dysregulated host response evolves remains unclear because few studies have followed patients at single-cell resolution. We profiled single-cell transcriptomes, cell-surface proteomes and T cell receptor repertoires of blood mononuclear cells with matched plasma proteomes across 305 samples from 71 adults with sepsis and 27 with sterile inflammation, from emergency department presentation to convalescence, plus 12 healthy controls. A CD14⁺ monocyte state with low MHC class II gene expression was expanded at presentation before contracting progressively, whereas dendritic cells and naive CD8⁺ T cells were low initially and recovered more slowly. A latent factor integrating these measurements was associated with illness severity and time of measurement, differing between survivors and non-survivors by day 3, but not at presentation. Because up to 73% of patients changed consensus sepsis endotype class within the first week, stratification and immunomodulation may require serial measurement.

## INTRODUCTION

Sepsis, defined as life-threatening organ dysfunction arising from a dysregulated host response to infection, causes an estimated 11 million deaths each year, yet no immunomodulatory therapy has reproducibly improved outcomes in patients^1,2^. One reason is that the dysregulated response is neither uniform nor static: across patients, it spans a spectrum from hyperinflammation to profound immune suppression; and within patients, these states evolve, and often coexist, over the hours to weeks of acute illness^3–5^. Hyperinflammation involves release of cytokines such as IL-6 and TNF-α, complement activation, and endothelial disruption^6–8^, whereas manifestations of immune suppression include lymphocyte depletion, impaired antigen presentation, and the emergence of dysfunctional or inhibitory myeloid populations^9–11^. The magnitude and resolution kinetics of these perturbations vary across patients, but their contribution to heterogeneity in clinical trajectories and outcomes is unknown.

Molecular profiling has made inroads into parsing this heterogeneity. Bulk blood transcriptomics has defined reproducible sepsis subtypes or “endotypes”^12–16^, among them the sepsis response signature (SRS) classification, which stratifies mortality risk and predicts response to corticosteroids^13,17^. Two recent consensus efforts have reconciled multiple endotyping schemas into three consensus transcriptomic subtypes^18^ and four classes defined by the degree of myeloid and lymphoid dysregulation^19^. Because bulk measurements average over mixed leukocytes, however, the cell types and transcriptional states that generate these endotypes are inferred rather than observed. A second limitation is temporal. The endotypes in current use were derived almost exclusively from single samples drawn on admission to an intensive care unit (ICU) rather than at first presentation to care^13,15,18,19^, placing the reference for many endotype definitions at least hours and up to several days later, and after the initial response to clinical management. Recent longitudinal analysis of the whole-blood transcriptome in sepsis has shown that immune-response trajectory and clinical stage are only partly aligned, so samples drawn at the same clinical timepoint may occupy different points along a shared immune trajectory^20^. Whether an endotype assignment reflects an enduring feature of a patient’s immune response or its state at the moment of sampling is unresolved, and establishing how rapidly endotype assignment changes within individual patients has been identified as a major research priority^18^.

Single-cell approaches can resolve the cell states that generate bulk signatures, but have been applied to sepsis only rarely^21–23^. We previously identified a CD14⁺ monocyte state, termed MS1, that expands in sepsis and has transcriptional hallmarks of monocytic myeloid-derived suppressor cells (M-MDSCs), including low HLA-DR and high expression of alarmins S100A8 and S100A9^23^. Suppression of T cell responses is the defining functional property of an MDSC^24^, and plasma from sepsis patients induces MS1-like cells from hematopoietic progenitors *in vitro* that suppress T cell proliferation^25^. Whole-blood single-cell profiling of sepsis samples has described a granulocytic counterpart to this state: IL1R2^+^ immature neutrophils, which arise through emergency granulopoiesis, carry transcriptional hallmarks of granulocytic MDSCs, and suppress T cell activation and proliferation^21^. These single-cell studies have almost uniformly been cross-sectional, with only scant assessment of immune responses at convalescence.

We therefore assembled a cohort of patients to longitudinally assess the circulating host immune response in sepsis as it evolves. Adults with sepsis or sterile inflammation (i.e., acute organ dysfunction without infection) were enrolled at first presentation to an Emergency Department (ED) rather than on ICU admission, and were sampled from presentation through the first week of treatment and into convalescence. Sterile inflammation was chosen as the comparator because such patients often receive the same empirical management as patients with sepsis, but have no microbial stimulus. The comparison therefore separates features of the immune response that require infection from those that accompany severe acute illness and its treatment. We performed CITE-seq (joint single-cell transcriptome and surface-protein measurement)^26^ with T cell receptor (TCR) sequencing on peripheral blood mononuclear cells (PBMCs) and targeted proteomics on matched plasma. This design allowed us to characterize the immune state of early sepsis before sustained intervention and to follow its evolution with treatment; to investigate whether the different multi-omic features reflect coordinated host responses; and to assess the temporal stability of, and cellular basis for, transcriptional endotype assignments within individual patients.

## RESULTS

### MS1 monocytes are expanded in sepsis at clinical presentation

We enrolled adults presenting to the ED at Massachusetts General Hospital with suspected sepsis and acute organ dysfunction, within 12 hours of ED arrival, and before more than 12 hours of antibiotic exposure (Methods). Blood was drawn at presentation (day 0), and on days 1 and 3; additional samples were obtained in a subset on day 7 and at convalescence (≥28 days after the index episode) (Fig. 1a). Three blinded physician-investigators adjudicated each participant as having sepsis or sterile inflammation. The final cohort comprised 71 patients with sepsis, 27 with sterile inflammation, and 12 healthy controls, contributing 305 samples (Fig. 1a); unavailable later samples primarily reflected discharge, death, or loss to follow-up. Healthy controls were sampled once and serve throughout as a cross-sectional reference, because circulating immune composition in healthy adults is stable within individuals over time and returns to baseline after acute challenge^27,28^.

**Figure 1.**
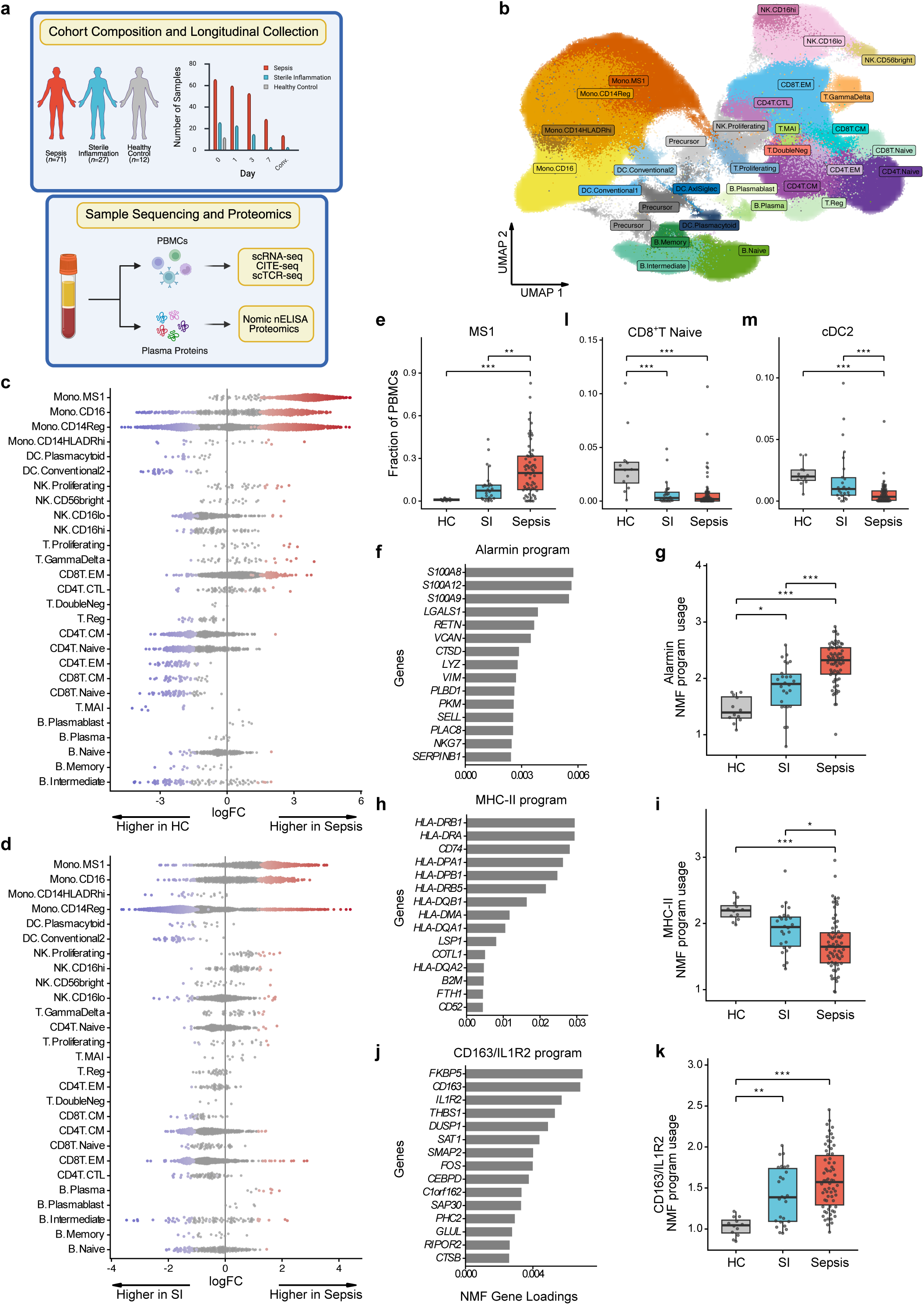
A multi-omic atlas identifies expansion of an MS1 monocyte state in sepsis at clinical presentation. **a**, Study design: cohort composition and serial sampling strategy, with peripheral blood mononuclear cells (PBMCs) profiled by cellular indexing of transcriptomes and epitopes by sequencing (CITE-seq) and single-cell T cell receptor (TCR) sequencing, and plasma by targeted proteomics; bar charts show samples per group at each timepoint. n = 71 patients with sepsis, 27 patients with sterile inflammation and 12 healthy controls, contributing 305 samples in total. **b**, Uniform Manifold Approximation and Projection (UMAP) embedding of integrated single cell transcriptome and surface proteome PBMC profiles, annotated by immune cell state. Cell states are annotated as lineage followed by transcriptional state, separated by a period; e.g. DC.Plasmacytoid denotes plasmacytoid dendritic cells; this convention is used in all panels that display state level labels. Mono denotes monocyte; Reg, regular; NK, natural killer; DC, dendritic cell; CM, central memory; EM, effector memory; CTL, cytotoxic T lymphocyte; MAIT, mucosal-associated invariant T cell. **c**,**d**, Neighborhood differential abundance at day 0, shown as beeswarms of per-neighborhood log fold-change grouped by lineage and ordered within lineage by significance, for sepsis versus healthy control (**c**) and sepsis versus sterile inflammation (**d**). Colored points are neighborhoods significant at a spatial false discovery rate below 0.1; positive values indicate enrichment in sepsis; non-significant neighborhoods are shown in grey. Cell states contributing fewer than three neighborhoods are omitted. **e**, Fraction of PBMCs in monocyte state 1 (MS1) by group at day 0. **f**, **h**, **j**, Top gene loadings of the alarmin (**f**), MHC class II (**h**), and CD163/IL1R2 (**j**) non-negative matrix factorization (NMF) gene programs. **g**, **i**, **k**, Sample-level activity of each gene program in monocytes by group at day 0. **l**, Naive CD8⁺ T cell fraction by group at day 0. **m**, Type 2 conventional dendritic cell (cDC2) fraction by group at day 0. In **e**, **g**, **i**, **k**, **l**, and **m**, boxes show median and interquartile range, whiskers extend to 1.5× the interquartile range, and points are individual samples. The groups are sepsis and sterile inflammation at day 0 and healthy controls; groups were compared by linear models adjusted for age and sex with Tukey-adjusted pairwise contrasts; * denotes *P_adj_* < 0.05, **, *P_adj_* < 0.01, ***, *P_adj_* < 0.001; *P_adj_* denotes a *P* value adjusted for multiple comparisons. SI denotes sterile inflammation; HC, healthy control.

Illness severity at presentation, measured by the Sequential Organ Failure Assessment (SOFA) score, did not differ between groups, and age distributions overlapped (Extended Data Fig. 1a,b); all cross-sectional comparisons were adjusted for age and sex. Among patients with sepsis, infections developed from the respiratory and urinary tracts, abdomen, endovascular sources and skin; 52% of patients were bacteremic (Extended Data Fig. 1c,d).

CITE-seq^26^ yielded 560,867 high-quality PBMC transcriptomes with paired surface-protein measurement, resolving 29 immune cell states across monocytes, dendritic cells (DCs), T cells, B cells and natural killer (NK) cells (Fig. 1b). Among monocytes, we recovered a CD14⁺ state with high expression of *S100A8*, *S100A9*, resistin (*RETN*), interleukin-1 receptor type 2 (*IL1R2*), and clusterin (*CLU*), as well as reduced *HLA-DRA* and *HLA-DRB1*, corresponding to the MS1 state previously associated with bacterial sepsis^23^.

Neighborhood-based differential abundance testing^29^ demonstrated that PBMC remodeling at presentation was dominated by expansion of MS1, which was enriched in sepsis relative to healthy controls (Fig. 1c; Supplementary Fig. 1a) and sterile inflammation (Fig. 1d; Supplementary Fig. 1b). Dendritic cell, B cell, and naive and central memory T cell neighborhoods were depleted, while cytotoxic and proliferating T cell neighborhoods were enriched relative to healthy controls (Fig. 1c, Supplementary Fig. 1a). At the sample level, the MS1 fraction was higher in sepsis than in healthy controls (*P_adj_* = 4.0 × 10⁻⁴; Fig. 1e) and higher than in sterile inflammation (*P_adj_* = 5.0 × 10⁻³; Fig. 1e), whereas neither regular CD14^+^ nor HLA-DR^hi^ CD14^+^ monocytes were expanded in sepsis (Supplementary Fig. 1c,d). Sepsis therefore shifts monocyte composition toward MS1 rather than expanding classical monocytes as a whole.

Cell-state labels are discrete, whereas transcriptional programs are graded and can be shared across states. We therefore used non-negative matrix factorization, an unsupervised method that recovers sets of genes co-expressed across cells (i.e., gene programs)^30^, and scored their activity at the sample level within immune-cell lineages (Methods; Supplementary Fig. 2). An alarmin-enriched gene program in monocytes (Fig. 1f) increased stepwise from healthy controls through sterile inflammation to sepsis (all pairwise contrasts significant; Fig. 1g). An MHC class II program (Fig. 1h) showed the reciprocal pattern and was reduced in sepsis relative to both comparators (Fig. 1i). A third program, loading on *CD163* and *IL1R2,* together with the glucocorticoid-responsive transcripts *FKBP5* and *THBS1* (Fig. 1j), was elevated in both sepsis and sterile inflammation relative to healthy controls (Fig. 1k). The alarmin and CD163/IL1R2 programs were correlated across samples (Spearman ρ = 0.71 for all samples, 0.68 within sepsis; Supplementary Fig. 1e), indicating that the two programs rise together rather than marking mutually exclusive groups of patients.

Outside the monocyte lineage, naive CD8^+^ T cells were reduced in both inflammatory groups relative to healthy controls (*P*_adj_ = 1.7 × 10⁻⁴ and 1.1 × 10⁻⁴), but did not differ between them (Fig. 1l). Conventional type 2 DCs were reduced in sepsis relative to both healthy controls (*P*_adj_ = 7.7 × 10⁻⁴) and sterile inflammation (*P*_adj_ = 4.2 × 10⁻⁴; Fig. 1m); cDC2 depletion, MS1 expansion and loss of the monocyte MHC class II program therefore distinguished sepsis from sterile inflammation of comparable severity at clinical presentation.

### MS1 contracts rapidly while lymphoid and antigen-presenting cell recovery is slow and incomplete at convalescence

To resolve the temporal structure of the immune changes from presentation through convalescence, we compared each sepsis timepoint with day 0 by neighborhood-level differential abundance testing. Median log fold-change (logFC) in MS1 representation relative to day 0 was −0.24 at day 1, −1.17 at day 3 (Fig. 2a,b), −1.76 at day 7, and −1.86 at convalescence (Extended Data Fig. 2), whereas regular and HLA-DR^hi^ CD14⁺ monocyte neighborhoods moved reciprocally, as did dendritic cell and memory T cell neighborhoods (Fig. 2a,b; Extended Data Fig. 2).

**Figure 2.**
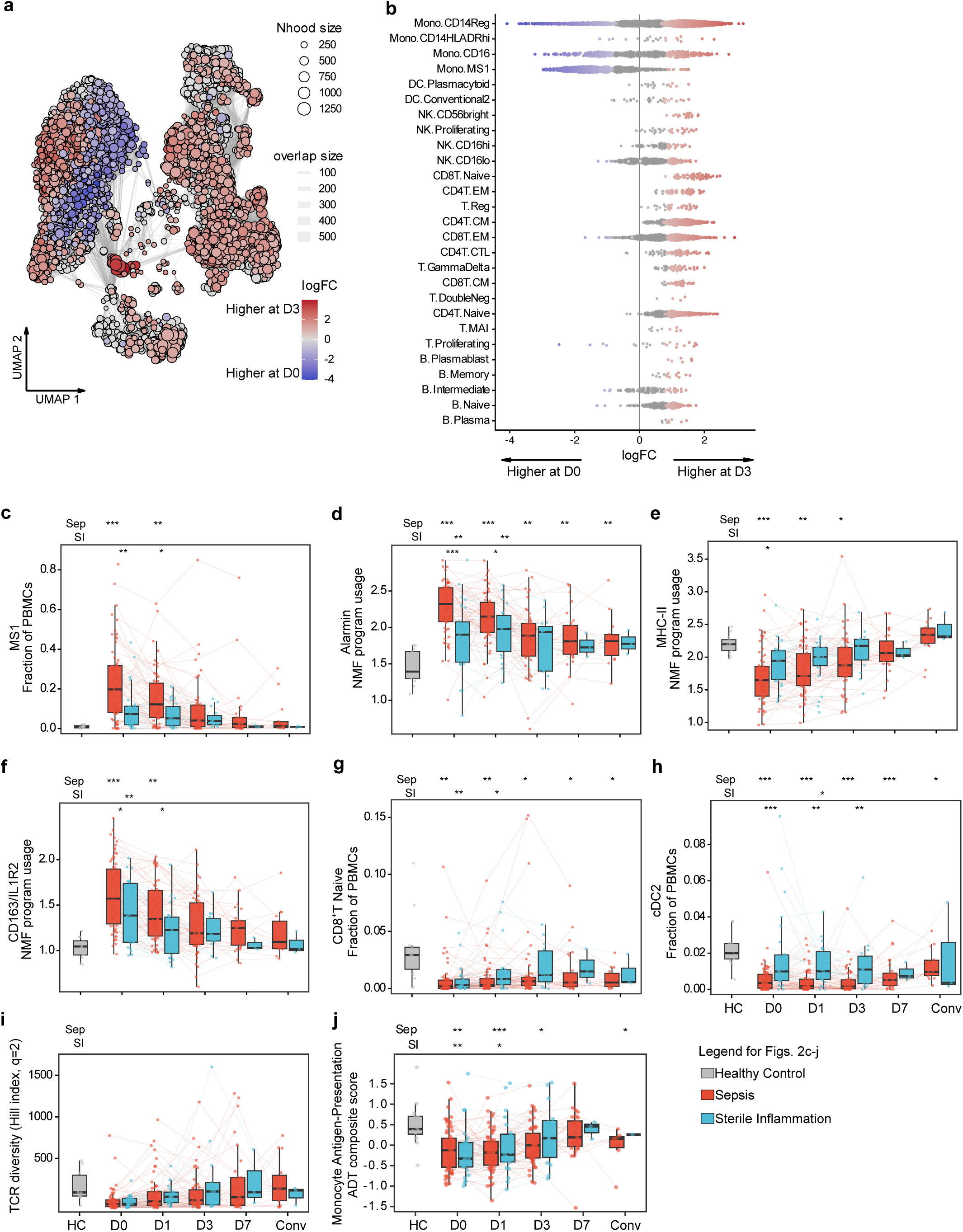
MS1 contracts rapidly while lymphoid and antigen-presenting cells recover slowly. **a,b**, Neighborhood differential abundance between day 3 and day 0 in patients with sepsis only, shown as the neighborhood graph projection (**a**) and a beeswarm of per-neighborhood log fold-change grouped by cell lineage (**b**). Negative log fold-changes (blue) indicate lower abundance at day 3 than at day 0 and positive values (red) higher abundance at day 3; node size in **a** is the neighborhood cell count and edge width the number of cells shared between neighborhoods. 4,399 neighborhoods were tested across paired day 0 and day 3 samples. **c–h**, Sample-level longitudinal trajectories in patients with sepsis and with sterile inflammation, with healthy controls shown once as a cross-sectional reference, for: MS1 fraction (**c**), and monocyte activity of the alarmin (**d**), MHC class II (**e**), and CD163/IL1R2 (**f**) gene programs; naive CD8⁺ T cell fraction (**g**), and cDC2 fraction (**h**). **i**, Longitudinal trajectory of T cell repertoire diversity (assessed by Hill number at order q = 2) estimated with log-transformed TCR sequencing depth as a covariate in the same three groups as c-h. **j**, Monocyte antigen-presentation cell surface-protein composite (CITE-seq antibody-derived tags [ADT] of HLA-DRA, HLA-A, and CD86, scored across monocyte states), longitudinal trajectory in the same three groups as c-h. For **c**-**j**: boxes show median, interquartile range and 1.5× the interquartile range, thin lines connect samples from the same patient, the upper symbol rows denote significance versus healthy controls for sepsis (“Sep”) and sterile inflammation (“SI”) at each timepoint, and in-plot symbols denote sepsis versus sterile inflammation; comparisons use linear mixed-effects models with a per-patient random effect adjusted for age and sex (* denotes *P_adj_*<0.05, ** *P_adj_*<0.01, *** *P_adj_*<0.001). Samples per group at each timepoint are shown in Fig. 1a. D0, D1, D3, and D7 denote days 0, 1, 3, and 7 after presentation; Conv, convalescence; HC, healthy control; SI, sterile inflammation; MS1, monocyte state 1; cDC2, type 2 conventional dendritic cell; TCR, T cell receptor; CITE-seq, cellular indexing of transcriptomes and epitopes by sequencing. Day 0 values are the same measurements shown cross-sectionally in Fig. 1; statistics here are from longitudinal mixed-effects models (Methods).

Sample-level trajectories were then compared between the two inflammatory groups, and each timepoint in each group was compared with the healthy-control reference; the direction of each principal contrast was preserved when any single control was omitted (Methods; Supplementary Table 1). In sepsis, the MS1 fraction was elevated relative to healthy controls at days 0 and 1 but was not significantly different by day 3 (Fig. 2c). Alarmin program activity decreased after presentation but remained above control levels through convalescence (Fig. 2d), MHC-II program activity returned to control levels by day 7 (Fig. 2e), and the CD163/IL1R2 program declined over the first week in both inflammatory groups (Fig. 2f). Recovery was not uniform: naive CD8⁺ T cells (Fig. 2g) and cDC2s (Fig. 2h) remained below control levels at every timepoint including convalescence, as did plasmacytoid DCs and memory B cells (Extended Data Figs. 3f and 4l).

Because fractional abundances are constrained to sum to one, we also estimated absolute abundances by scaling each state within its lineage to the matched clinical monocyte or lymphocyte count (Methods). These were broadly concordant with the fractional trajectories (Extended Data Figs. 5 and 6). In sepsis, absolute MS1 numbers fell from day 0 to day 3 and were stable thereafter (Extended Data Fig. 5a).

In an independent whole-blood single-cell sepsis cohort^21^, reclustering CD14^+^ monocytes resolved an MS1 state that the original annotation had not separated, allowing it to be compared with the IL1R2^+^ immature neutrophil state defined in that study as its putative granulocytic equivalent. In nine sepsis patients with paired acute and convalescent samples, both cell states contracted with recovery, down to healthy control levels (*P*_adj_ = 0.011 for IL1R2^+^ immature neutrophils and 0.035 for MS1; Extended Data Fig. 7a,b). Type 2 conventional dendritic cells and naive CD4^+^ T, CD8^+^ T, and B cells had not fully recovered to healthy control levels at convalescence, but the differences did not reach statistical significance (Extended Data Fig. 7c-f).

To determine whether the changes in T cell composition in our cohort were accompanied by a change in the clonal composition of the repertoire, we used paired transcriptome and TCR sequences (Methods). Expanded clonotypes concentrated in cytotoxic and activated states (Supplementary Fig. 3a,b). Both inflammatory groups contained more expanded clones than healthy controls, with similar clone-size distributions (Supplementary Fig. 3c), and pairwise repertoire overlap between patients was minimal (Supplementary Fig. 4). We quantified diversity using the Hill number of order q = 2, which is sensitive to clonal expansion and the least sensitive of the Hill orders to the incomplete sampling inherent in single-cell TCR profiling^31^. Diversity of the total T cell repertoire was directionally lower in sepsis at presentation than in healthy controls, but the difference did not reach significance at any timepoint (Fig. 2i). This result was adjusted for sequencing depth and was unchanged when every sample was rarefied to a common depth; individual T cell subsets contained too few TCR-bearing cells for diversity to be estimated reliably (Supplementary Table 2).

Paired surface-protein measurement corroborated the monocyte antigen-presentation transcriptional findings. A composite of markers (HLA-DRA, HLA-A and CD86; Methods) was reduced relative to healthy controls in sepsis through day 3, and was again lower at convalescence (Fig. 2j). T cell homeostasis, T cell checkpoint and dendritic cell antigen-presentation composites did not differ from controls at any timepoint (Extended Data Fig. 8). Together, these results describe asynchronous recovery from acute sepsis: MS1 monocytes expand early and contract within days, whereas dendritic cell and naive CD8^+^ T cell populations recover more gradually and are not fully restored at clinical convalescence.

### Plasma proteins associated with the monocyte response resolve on different timescales

The plasma proteome reflects dimensions of the host response that are not directly captured by circulating immune cells, including cytokine and chemokine release, endothelial cell dysfunction, and end-organ injury. We therefore profiled matched plasma with a targeted antibody-based immunoassay (nELISA) that quantifies 264 secreted proteins, primarily cytokines, chemokines, growth factors and soluble receptors, as absolute concentrations calibrated against reference standards; 117 proteins passed a prespecified detectability filter and were analyzed on a log₁₀ scale (Methods). Across sepsis samples from all timepoints, we identified multiple proteins that were positively associated with MS1 abundance and with the alarmin and CD163/IL1R2 gene programs, and inversely associated with the MHC-II gene program. The strongest associations were with acute-phase proteins, interleukins and their receptors, chemokines, TNF receptors and TGFβ-family proteins (Fig. 3a; full analyte set in Extended Data Fig. 9a). Correlation-based clustering (Methods) yielded four protein modules (Supplementary Table 3), of which module 1 comprised many of these proteins. Because MS1 monocytes are thought to arise through cytokine-driven emergency myelopoiesis^25,32^, we next examined the longitudinal trajectories of plasma proteins linked to that process (IL-6, IL-8) and to the MS1 state itself (resistin, CHI3L1). IL-6 and IL-8 were elevated at day 0 in sepsis relative to both sterile inflammation and healthy controls, then declined over the acute course (Fig. 3b,c), whereas CHI3L1 and resistin were more persistently abnormal in sepsis, elevated relative to sterile inflammation early and above control levels through convalescence (Fig. 3d,e).

**Figure 3.**
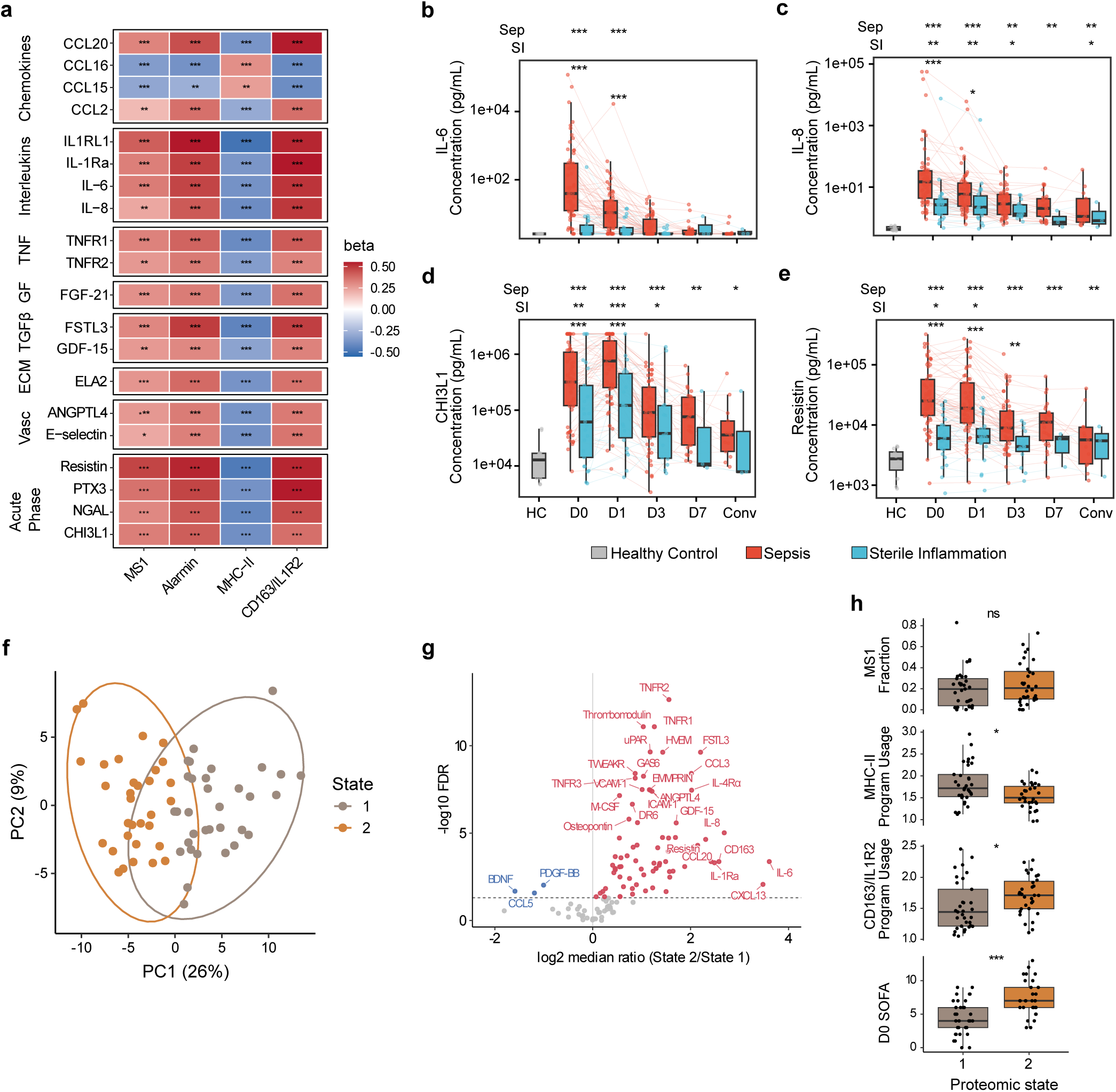
Plasma proteins associated with the monocyte response resolve on different timescales. **a,** Associations between plasma protein concentrations and MS1 fractional abundance as well as the alarmin, MHC class II, and CD163/IL1R2 gene programs, estimated across sepsis samples from all timepoints with a per-patient random intercept adjusted for age and sex. The 20 proteins with the largest mean absolute coefficient across the four features are shown. Proteins are grouped by functional class: chemokines (Chemokines); interleukins and their receptors (Interleukins); TNF superfamily proteins (TNF); growth factors (GF); TGFβ and BMP family proteins (TGFβ); proteases and extracellular matrix proteins (ECM); endothelial and adhesion proteins (Vasc); and acute-phase proteins (Acute Phase). Color shows the standardized coefficient; asterisks denote the Benjamini–Hochberg false discovery rate (FDR), where *< 0.05, **< 0.01, and ***< 0.001. **b-e,** Longitudinal plasma concentrations of IL-6 (**b**), IL-8 (**c**), CHI3L1 (**d**), and resistin (**e**) in patients with sepsis and with sterile inflammation, with healthy controls shown once as a cross-sectional reference; boxes show median, interquartile range and 1.5× the interquartile range. Thin lines connect samples from the same patient, the upper symbol rows denote significance versus healthy controls for sepsis (“Sep”) and sterile inflammation (“SI”) at each timepoint, and in-plot symbols denote sepsis versus sterile inflammation; comparisons used linear mixed-effects models with a per-patient random effect adjusted for age and sex (* denotes *P_adj_*<0.05, ** *P_adj_*<0.01, *** *P_adj_*<0.001). D0, D1, D3, and D7 denote days 0, 1, 3, and 7 after presentation; Conv, convalescence; HC, healthy control; SI, sterile inflammation. **f-h,** Plasma proteomic states, defined and compared in day 0 samples from patients with sepsis. **f,** Day 0 sepsis samples plotted on the first two principal components of the 117 plasma proteins passing the prespecified detectability filter (Methods) and colored by k-means proteomic state (k = 2); ellipses enclose 95% of a bivariate normal distribution fitted to each state. States were assigned on the first 14 principal components, which together explained 75% of the variance, so the two states can appear to overlap in this two-dimensional projection. **g,** Volcano plot of plasma protein abundance in proteomic state 2 relative to state 1 among day 0 sepsis samples, with the log₂ ratio of state medians and the −log₁₀ false discovery rate (two-sided Wilcoxon rank-sum test, Benjamini–Hochberg-adjusted); proteins higher in state 2 are shown in red and proteins lower in state 2 in blue. **h,** MS1 fraction, monocyte MHC class II and CD163/IL1R2 program activity, and day 0 SOFA score by proteomic state in day 0 sepsis samples; boxes show median, interquartile range and 1.5× the interquartile range (two-sided Wilcoxon rank-sum test; * denotes *P*<0.05, **, *P*<0.01, *** *P*<0.001, ns, not significant). n = 65 day 0 sepsis samples with matched plasma, 32 in state 2 and 33 in state 1. MS1 denotes monocyte state 1; PBMC, peripheral blood mononuclear cell.

To define proteomic states at presentation, and before treatment had substantially reshaped the response, we performed unsupervised clustering of day 0 sepsis samples on the 117 proteins; this identified two states (k-means on principal components, k = 2 by silhouette; Fig. 3f). Later samples were assigned to these states without contributing to their definition. Most differentially abundant proteins were higher in state 2, including soluble TNF-receptor superfamily members (TNFR1, TNFR2, HVEM, DR6), the interleukins IL-6, IL-8 and IL-1Ra, and endothelial proteins, whereas only BDNF, CCL5 and PDGF-BB were lower (Fig. 3g). State 2 had lower MHC-II program activity (*P* = 0.019) and higher CD163/IL1R2 program activity (*P* = 0.023), whereas MS1 abundance did not differ (Fig. 3h). We also compared the two states on two clinical outcomes that were applied to other measures in this study: illness severity at presentation and in-hospital mortality (Methods). State 2 had a higher day 0 SOFA score (*P* = 1.5 × 10⁻⁴; Fig. 3h), and in-hospital mortality was non-significantly higher in state 2 (Fisher’s exact test *P* = 0.061; Extended Data Fig. 9b).

### A PBMC-derived continuous endotype score captures heterogeneous myeloid and lymphoid dysfunction in sepsis

Having characterized the cellular, transcriptional and plasma features of the sepsis response, we next asked how they relate to transcriptional signatures used to define endotypes. Established sepsis endotypes are defined on whole blood, in which mononuclear and granulocyte signals are mixed and the relative cellular contributions to different endotype signatures cannot be resolved. Because granulocytes, the most abundant circulating leukocytes, are excluded from PBMC studies such as this one, it is uncertain how much of a whole-blood endotype signature can be recovered from mononuclear cells alone. To test this directly, we reanalyzed a published single-cell sepsis cohort^21^ profiled from whole blood, in which granulocytes were retained and each sample carries an SRS assignment (SRS1 versus non-SRS1) together with the quantitative SRS score (SRSq) from which that assignment was derived. In that cohort, we aggregated expression across cells in each sample to form pseudobulk profiles from all leukocytes (whole-blood pseudobulk) and from mononuclear cells alone (mononuclear pseudobulk), and tested recovery of the reference labels in three ways: the supervised classifier SepstratifieR^33^ applied to each, and expression of the 19-gene SRS signature^33^ in the mononuclear pseudobulk, hereafter the PBMC SRS score. The PBMC SRS score correlated closely with SRSq (Spearman ρ = 0.68, *P* < 0.001) and separated SRS1 from non-SRS1 samples with an area under the receiver operating characteristic curve (AUROC) of 0.89 (Fig. 4a, Supplementary Table 4). Applied to the same samples with the same 19 genes, SepstratifieR reached an AUROC of 0.94 on whole blood but only 0.64 on the mononuclear pseudobulk. We therefore carried the PBMC SRS score forward as a continuous measure, thus preserving graded differences between samples and permitting direct quantitative association with the cellular and clinical features measured in the same samples.

**Figure 4.**
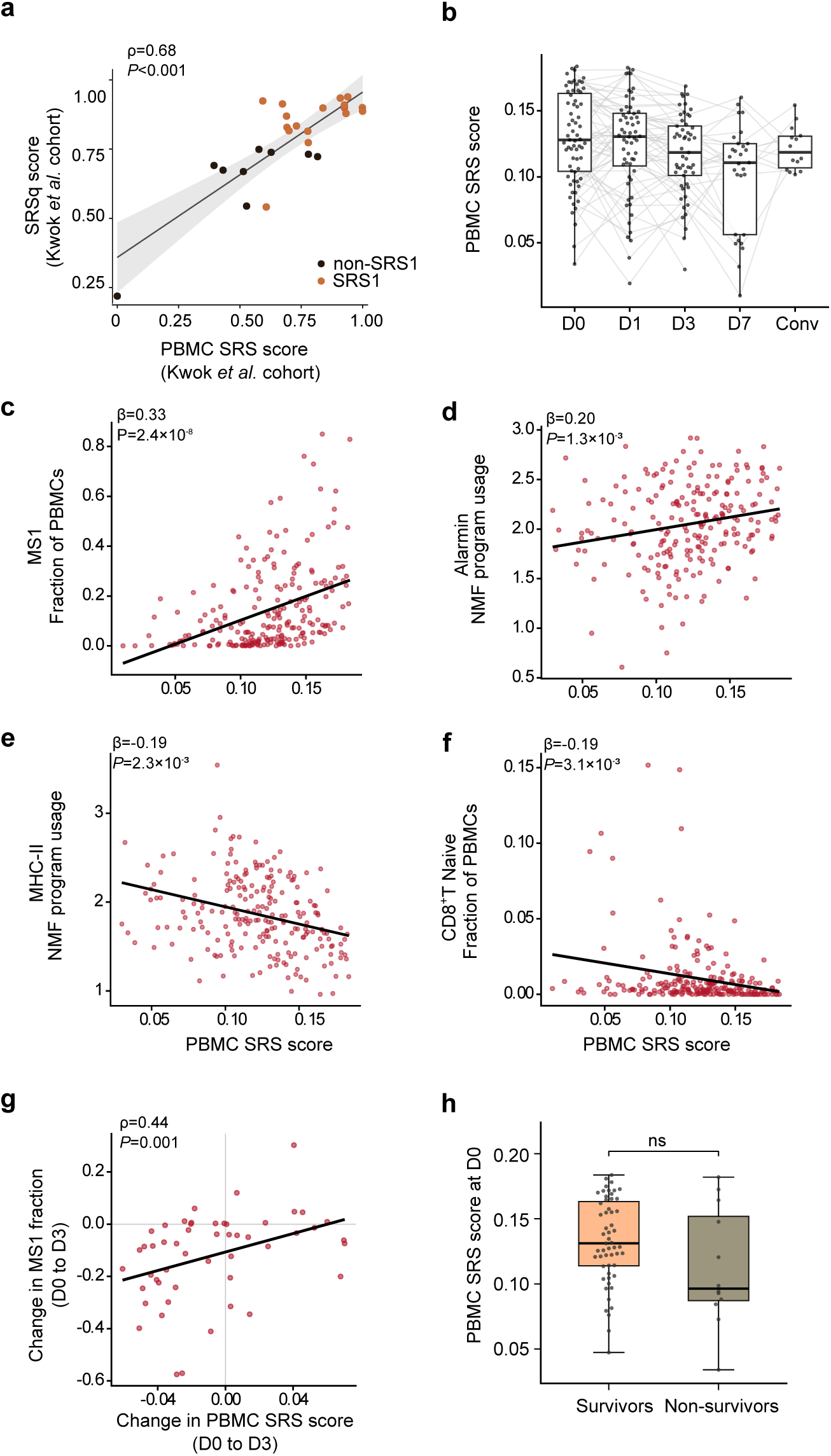
A PBMC-derived SRS signature score is associated with MS1 expansion, loss of the monocyte MHC class II program and depletion of naive CD8⁺ T cells in sepsis. **a**, Peripheral blood mononuclear cell (PBMC) sepsis response signature (SRS) score benchmarked against a matched whole-blood quantitative SRS score (SRSq) in an independent single-cell reference cohort^21^ (n = 26 samples; 16 SRS1, 10 non-SRS1), colored by reference class; the line is a linear fit with its 95% confidence band, the Spearman ρ and *P* value are annotated. **b**, Longitudinal trajectory of the PBMC SRS signature score in sepsis samples; boxes show median, interquartile range and 1.5× interquartile range, with thin lines linking samples from the same patient. **c–f**, Associations of the PBMC SRS score with MS1 abundance (**c**), the alarmin program (**d**), the MHC class II program (**e**) and naive CD8⁺ T cell abundance (**f**) across sepsis samples; annotations show the standardized coefficient from a linear mixed-effects model with a per-patient random intercept adjusted for sampling day. **g**, Spearman correlation of the within-patient change in MS1 fraction with the change in PBMC SRS score between presentation and day 3; each point is one sepsis patient. **h**, PBMC SRS score at day 0 in sepsis survivors and those who died in the hospital (two-sided Wilcoxon rank-sum test; ns denotes not significant). D0, D1, D3, and D7 denote days 0, 1, 3, and 7 after presentation; Conv, convalescence.

Across sepsis samples, the PBMC SRS score was higher at presentation and declined thereafter (Fig. 4b), and was associated with greater MS1 abundance (β = 0.33, *P* = 2.4 × 10⁻⁸), higher alarmin program activity (β = 0.20, *P* = 1.3 × 10⁻³), lower MHC-II program activity (β = −0.19, *P* = 2.3 × 10⁻³) and fewer naive CD8⁺ T cells (β = −0.19, *P* = 3.1 × 10⁻³), each estimated in a mixed-effects model that included sampling day as a covariate (Fig. 4c–f; Methods). Because MS1 abundance was the strongest of these correlates, we asked whether the two measures also changed congruently within individuals. Between presentation and day 3, the first timepoint at which the MS1 fraction no longer differed from healthy controls (Fig. 2c), the change in PBMC SRS score correlated with the change in MS1 abundance (ρ = 0.44, *P* = 0.001; Fig. 4g).

We next asked whether the PBMC SRS score was associated with clinical severity and outcome. Because the score was measured serially, two assessments were added to those applied to the proteomic states, for this and subsequent immune measures (Methods). First, sepsis survivors and non-survivors were compared at each timepoint separately; second, the within-patient change in the measure between presentation and day 3 was compared with the change in SOFA score over the same interval. The PBMC SRS score showed none of these associations. At presentation it was associated with neither SOFA score (Extended Data Fig. 10a) nor in-hospital mortality (Fig. 4h); it did not differ between survivors and non-survivors at any later timepoint (Extended Data Fig. 10b), and its change between presentation and day 3 was unrelated to the change in SOFA score (Extended Data Fig. 10c).

### A latent factor integrating multi-omic inputs differs between sepsis survivors and non-survivors by day 3

Multiple sample-level features in the preceding analyses were correlated with one another, raising the question of whether they represent parallel findings or a coordinated process. We therefore applied multi-omics factor analysis^34^ (a generalization of principal component analysis for multi-omics data), which has previously been used to integrate cross-sectional data in sepsis^21,22^, to all the sepsis samples. This method summarizes the variation shared across its inputs as a small number of latent factors, each a weighted combination of measurements that vary together across samples. Here, the model integrated 39 sample-level measurements of five kinds: PBMC cell state abundances, monocyte and T cell gene programs, TCR diversity metrics, surface protein composites and plasma protein measurements. Neither sampling day nor any clinical variable was included as a model input. We then examined how each of the eight fitted factors related to sampling day, illness severity, the PBMC SRS score and in-hospital mortality (Supplementary Table 5). One factor (Factor 4) was associated with all four after adjustment across factors, and had the strongest association with sampling day. The same factor had the greatest correlation with prespecified cellular features of the acute response characterized above (Supplementary Table 5; Methods), and was selected for further analysis.

Factor 4 was highest at presentation and declined over time (ρ = −0.43, *P* = 1.4 × 10⁻¹¹; Fig. 5a). Among the features tested (Fig. 5b, Supplementary Table 6), it loaded principally on the alarmin program (β = 0.73), the CD163/IL1R2 program (β = 0.70), MS1 abundance (β = 0.69) and, inversely, the MHC-II program (β = −0.68; all *P_adj_*<0.001). All DC states were inversely associated (β = −0.38 to −0.15). Higher factor values were associated with higher PBMC SRS scores (β = 0.18, *P* = 0.002; Fig. 5c); they were also associated with higher SOFA scores on the same day (β = 0.21, *P* = 0.001; Fig. 5d).

**Figure 5.**
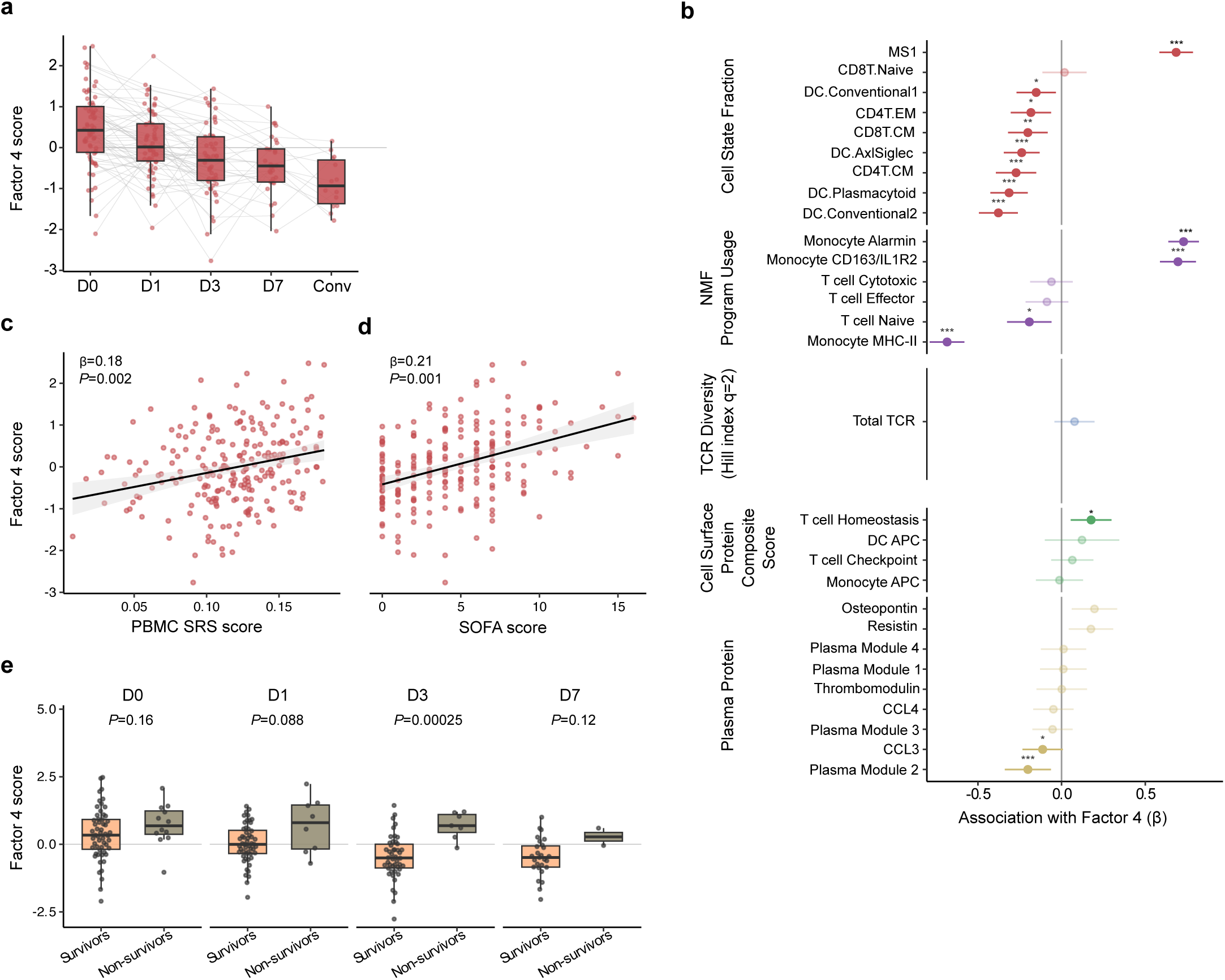
A multi-omics latent factor integrating cellular, repertoire, surface-protein and plasma measurements differs between sepsis survivors and non-survivors by day 3. **a,** Values of the selected latent factor across time for sepsis patients, derived from a multi-omics factor analysis (MOFA) of peripheral blood mononuclear cell (PBMC) cell state abundances, non-negative matrix factorization (NMF) gene program scores, T cell receptor (TCR) diversity features, cell surface protein composites and plasma protein measurements. Each point is one sample; grey lines connect serial samples from the same individual; boxplots show the median and interquartile range. **b,** Associations of the selected latent factor (Factor 4) with the input features, estimated by linear mixed-effects models (factor value regressed on each scaled feature with a per-patient random intercept). Points are point estimates and horizontal bars 95% confidence intervals; features significant after Benjamini–Hochberg adjustment across all features are shown in full color and non-significant features are faded. * denotes *P_adj_* < 0.05, **, *P_adj_* < 0.01, and ***, *P_adj_* < 0.001. **c**,**d**, Association of Factor 4 with the PBMC SRS score (**c**) and the day-matched SOFA score (**d**) across all sepsis samples; annotations show the standardized coefficient and *P* value from a linear mixed-effects model with a per-patient random intercept adjusted for sampling day. **e,** Factor value in survivors and in patients who died in the hospital, at each timepoint separately; boxes show median and interquartile range, points are individual samples, and annotations show the two-sided Wilcoxon rank-sum *P* value. n = 55, 52, 46 and 27 survivors and 12, 8, 7 and 2 non-survivors at D0, D1, D3, and D7, respectively. D0, D1, D3, and D7 denote days 0, 1, 3, and 7 after presentation; Conv, convalescence; SOFA, Sequential Organ Failure Assessment; SRS, sepsis response signature.

The relationship between Factor 4 and sepsis survival was time-dependent. At presentation, factor values did not differ between patients who died in hospital and survivors, whereas by day 3 they did (*P* = 2.5 × 10⁻⁴, Fig. 5e). Across all timepoints, death was associated with a higher factor value after adjustment for day (β = 0.59, *P* = 4.9 × 10⁻³). The mean factor across days 0 to 3 was higher in those who died (*P* = 0.036, Supplementary Fig. 5a), but the estimated rate of change between day 0 and day 3 did not differ by outcome (Supplementary Fig. 5b). Thus, the state reached by day 3, but not the state at presentation or the rate of change between them, was associated with survival.

We then assessed Factor 4 and MS1 abundance against the same clinical associations applied to the PBMC SRS score, so that a single cell state, a transcriptional signature, and an integration of all measured modalities could be compared directly (Extended Data Fig. 10). Unlike its association with outcome by day 3 (Fig. 5e), Factor 4 was not correlated with SOFA score at presentation (Extended Data Fig. 10d) and its change between presentation and day 3 was unrelated to the change in SOFA score (Extended Data Fig. 10f). MS1 abundance showed a different pattern: at presentation it correlated with SOFA score (ρ = 0.38, *P* = 0.002; Extended Data Fig. 10g) and its change between presentation and day 3 correlated with the change in SOFA score (ρ = 0.39, *P* = 0.005; Extended Data Fig. 10i), yet it did not differ between sepsis survivors and non-survivors at any timepoint (Extended Data Fig. 10h). Association with clinical severity and association with outcome are therefore separable properties, and improvement in SOFA score is an incomplete correlate for resolution of the broader immune response.

### Consensus endotype assignments change frequently during clinical management of sepsis

If the immune response resolves in ways that clinical assessment by SOFA score and similar metrics does not reflect, then an endotype assignment based on a single early sample may represent that patient’s host response for only a limited period of the illness. We therefore used the serial design of this cohort to assess whether endotype assignments remain stable within patients during standard clinical management of sepsis. We focused on the recent consensus frameworks: the consensus transcriptomic subtypes (CTS1–3)¹⁸ and the human immune dysregulation evaluation framework (Hi-DEF)¹⁹. Both were applied to every sample, with the plasma proteomic states described earlier used as a comparator derived in plasma rather than cells.

First, we determined how endotype assignment mapped onto cell state abundances and gene program activity. Among CTS endotypes, CTS1 had the highest MS1 abundance (threefold that of CTS3 and 24-fold that of CTS2; Fig. 6a) and alarmin program activity (Fig. 6b) as well as the fewest naive CD8⁺ T cells (Fig. 6d), while CTS2 had the lowest MS1 abundance and the highest MHC class II program activity (Fig. 6c). Within the Hi-DEF framework, samples were assigned almost entirely to the balanced and lymphoid dysregulation classes, with only rare assignment to system-wide dysregulation and none to isolated myeloid dysregulation, likely because the framework’s myeloid score draws largely on neutrophil-expressed genes^19^, which PBMCs do not capture. Samples in the lymphoid dysregulation class had eightfold higher MS1 abundance than those in the balanced class, with higher alarmin and lower MHC class II program activity, fewer naive CD8⁺ T cells and less than half the T cell repertoire diversity (Fig. 6a-e, lower panels). Factor 4 was highest in CTS1 and in the lymphoid dysregulation class (Fig. 6f). Thus, two classifiers built on different gene sets, and a factor derived without reference to either, rank their classes concordantly by MS1 expansion, loss of the monocyte MHC class II program and depletion of T cells.

**Figure 6.**
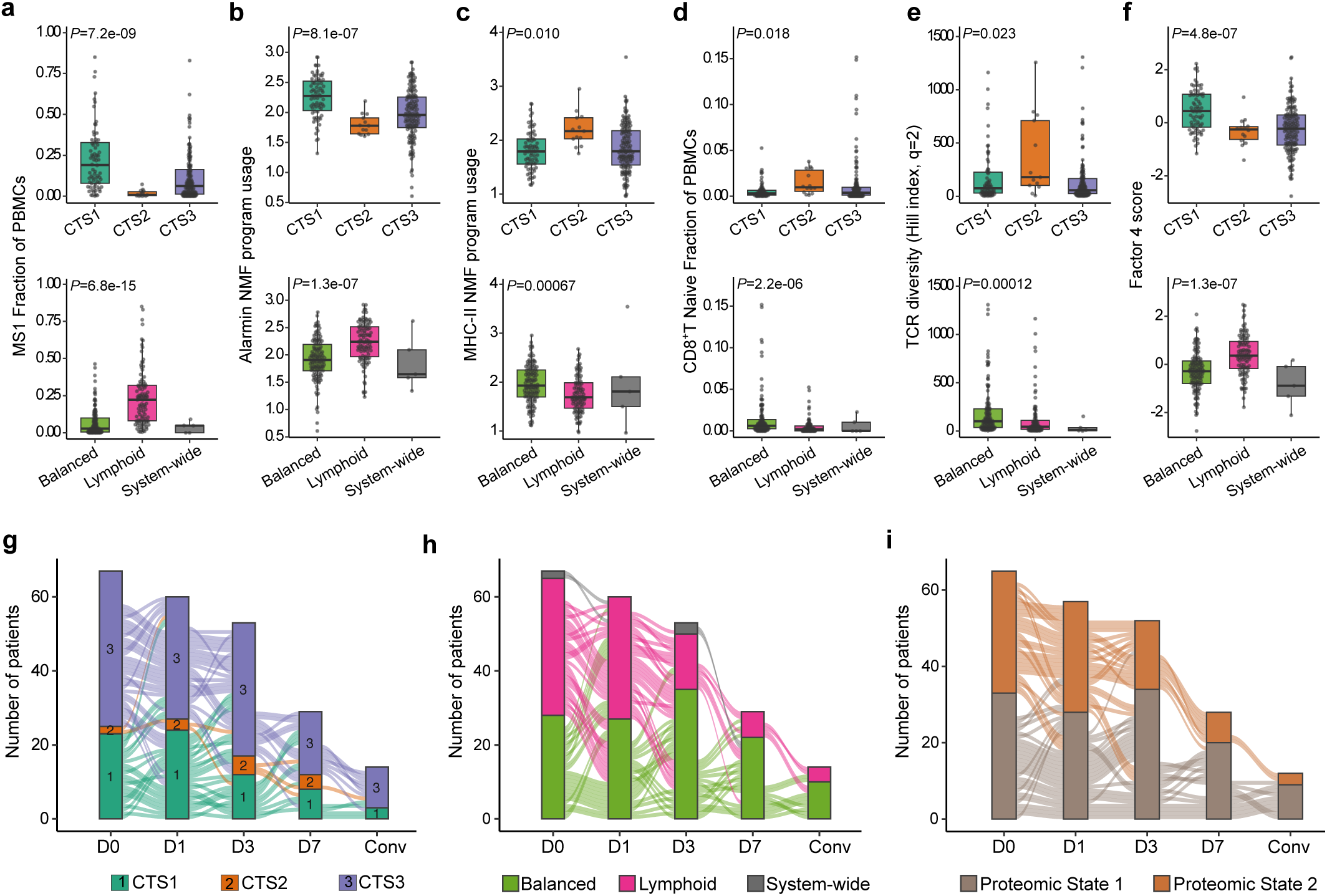
Consensus endotype assignments change over time. **a-f**, MS1 fraction (**a**), monocyte alarmin (**b**) and MHC class II (**c**) program activity, naive CD8⁺ T cell fraction (**d**), T cell receptor (TCR) diversity (**e**) and Factor 4 score (**f**) across consensus transcriptomic subtypes (CTS1–3; upper panels) and human immune dysregulation evaluation framework (Hi-DEF) classes (lower panels), in samples from patients with sepsis; boxes show median and interquartile range, points are samples, and annotations show Kruskal–Wallis *P* values across classes, Benjamini–Hochberg adjusted across the six features shown within each framework. Balanced, Lymphoid and System-wide denote the Hi-DEF balanced, lymphoid dysregulation and system-wide dysregulation classes; no sample was assigned to the myeloid dysregulation class. **g**-**i**, Alluvial diagrams of endotype assignment in patients with sepsis across days 0, 1, 3, 7 and convalescence, for CTS (**g**), Hi-DEF (**h**), and the plasma proteomic state (**i**). Each ribbon is one patient and each stratum one class at one timepoint; stratum height and ribbon width are numbers of samples, and ribbons are colored by the class the patient occupied at the preceding timepoint, so a ribbon that changes color is a patient who changed class. Totals fall over time because not every patient contributed every sample. D0, D1, D3, and D7 denote days 0, 1, 3, and 7 after presentation; Conv, convalescence.

Endotype assignments changed frequently over time. Among the 62 patients with sepsis contributing more than one sample, 53% changed CTS (Fig. 6g), and 73% changed Hi-DEF class (Fig. 6h) during the first week, whereas 30% changed plasma proteomic state over the same interval (Fig. 6i). Between presentation and day 3, movement out of CTS1, the lymphoid dysregulation class and plasma state 2 outnumbered movement into them: 12 versus 3 for CTS1 (*P* = 0.035), 21 versus 8 for lymphoid dysregulation (*P* = 0.024) and 10 versus 3 for plasma state 2 (*P* = 0.09). This asymmetry suggests that the frameworks summarize a resolving biological dysfunction rather than random fluctuation.

## DISCUSSION

Sepsis is characterized by the coexistence of inflammatory injury and immune dysfunction, yet the cellular and temporal organization of these processes has remained incompletely resolved, in part because most human studies have been cross-sectional. By profiling blood mononuclear cells and matched plasma from sepsis presentation through the first week of in-hospital management and into convalescence, we find a host response whose components resolve on different timescales during sepsis: expansion of the MS1 monocyte state and loss of monocyte MHC class II gene expression resolve within days, whereas dendritic cell and T cell populations recover more slowly and remain below healthy control levels at convalescence. Measurements of the host response therefore yield different results, including shifts in endotype assignments, depending on when samples are obtained.

Our findings are consistent with MS1 as a product of emergency myelopoiesis, the demand-adapted expansion of myeloid output that is induced acutely and subsides as the initiating insult is brought under control^35^; here, the abundance of MS1 cells in blood returned to control levels within the first days of treatment, before clinical recovery was complete. Plasma mediators tied to the same myeloid response divided along fast and slow kinetics: IL-6 and IL-8 were highest at presentation and decreased over a few days, whereas resistin, encoded by one of the genes that define MS1, remained above control levels into convalescence, after the cells themselves had contracted. Restoration of adaptive and antigen-presenting populations proceeded more slowly and incompletely. Naive CD8⁺ T cells, cDC2s, plasmacytoid DCs and memory B cells remained below healthy-control levels at convalescence. These findings imply that clinical convalescence may include a period of incomplete immune recovery, providing additional context for the delayed, immunosuppressed phase of sepsis that is associated with increased microbiological burden and late mortality^9,36^. While MS1 shares transcriptional features with monocytic MDSCs, whose defining function is suppression of T cell responses^24^, and MS1-like cells differentiated from progenitors in sepsis plasma suppress T cell proliferation^25^, whether MS1 contributes to the acute depletion of T cells in sepsis remains to be tested.

A latent host factor summarizing the multimodal measurements captured the temporal structure of the host response in sepsis, and its relationship to clinical outcome depended on when it was measured: the factor did not differ between survivors and non-survivors at presentation but did so by day 3. Candidate markers of immune suppression have demonstrated similar patterns across multiple cohorts: monocyte HLA-DR and absolute lymphocyte count were uninformative on day 1 but predictive of mortality in sepsis by day 3 or 4^37–40^. That the same pattern emerges from a multimodal factor derived empirically suggests that the timing is a property of the host response itself rather than of any particular assay. Endotype assignments showed the same dependence on timing. A comparable time-dependence has recently been described in the whole-blood transcriptome, in which patients with sepsis progress at different rates through three successive immune states, so that the clinical stage of illness is an incomplete indicator of a patient’s position along their immune trajectory^20^. Our cohort identifies monocyte gene programs as the main source of this variation within mononuclear blood cells, with patients moving over time out of the classes characterized by high MS1 abundance, high alarmin and low MHC-II program activity, and depletion of naive CD8⁺ T cells. Similar changes over time have been reported for sepsis plasma proteomic subphenotypes^41^ and for subtypes derived from routine clinical and laboratory variables^42^.

This study has several limitations. Blood profiling was restricted to PBMCs, and granulocytes were captured only through external data. Although this represents one of the largest single-cell studies of sepsis to date and, to our knowledge, the most extensively sampled longitudinally, the cohort was from a single center, and deaths were few. Treatment was not protocolized, by design, to capture real-world variation in sepsis management. Convalescent samples were obtained at heterogeneous times and were limited in number, which constrains conclusions about the timing and completeness of immune reconstitution. Both consensus endotype classifiers we evaluated were derived on whole blood but applied here to mononuclear cells, so the assignments and the switching frequencies describe the mononuclear fraction and cannot be assumed to represent whole-blood endotypes in the same patients; concordance between the two remains to be tested. In addition, the associations reported throughout this study, including those with illness severity and mortality, are observational and do not establish causal or mechanistic relationships.

In summary, longitudinal multimodal profiling frames the heterogeneity of sepsis as variation in position and rate of resolution along a trajectory of the host response, spanning emergency myelopoiesis and suppressive myeloid expansion, loss of monocyte antigen presentation, and slower recovery of adaptive immunity. As such, the timing of immune assessment, and of any immunomodulatory intervention in sepsis, must be considered alongside its target. Classifiers derived at ICU admission may therefore not carry the same meaning at first presentation to care, where the earliest treatment decisions are made. Enrichment of a clinical trial on a presentation sample may randomize patients to a class they have already left by the time the drug is given, so stratification may need to be established when treatment starts rather than at enrollment. Finally, because the association between the integrated host response and survival emerged only after the first few days, serial measurement may be more informative than a single baseline draw, both for prognosis and as a trial endpoint.

## Data Availability

All data produced in the present study are available upon reasonable request to the authors

## EXTENDED DATA FIGURES

**Extended Data Figure 1.**
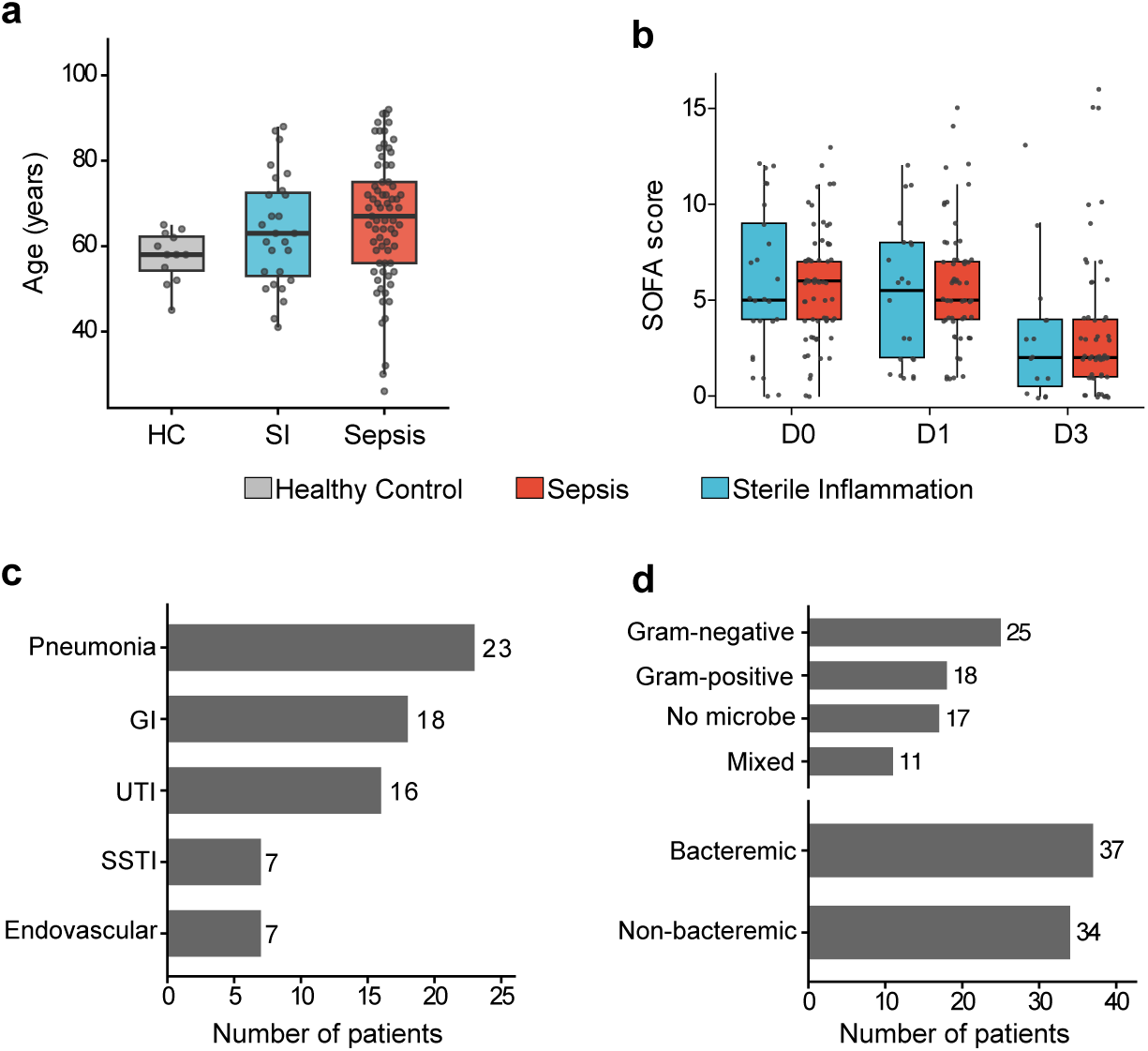
Cohort clinical characteristics. **a**, Age distribution by group. **b**, Sequential Organ Failure Assessment (SOFA) scores over time for the sepsis and sterile inflammation groups. **c**,**d**, Patients with sepsis stratified by source of infection (**c),** and Gram-stain category of microbial etiology and bacteremia status (**d)**. In **a** and **b**, boxes show median and interquartile range, whiskers extend to 1.5× the interquartile range, and points are individual patients (**a)** or samples (**b**). n = 71 patients with sepsis, 27 patients with sterile inflammation and 12 healthy controls. D0, D1, and D3 denote days 0, 1, and 3 after presentation; HC, healthy control; SI, sterile inflammation; GI, gastrointestinal; UTI, urinary tract infection; SSTI, skin and soft-tissue infection.

**Extended Data Figure 2.**
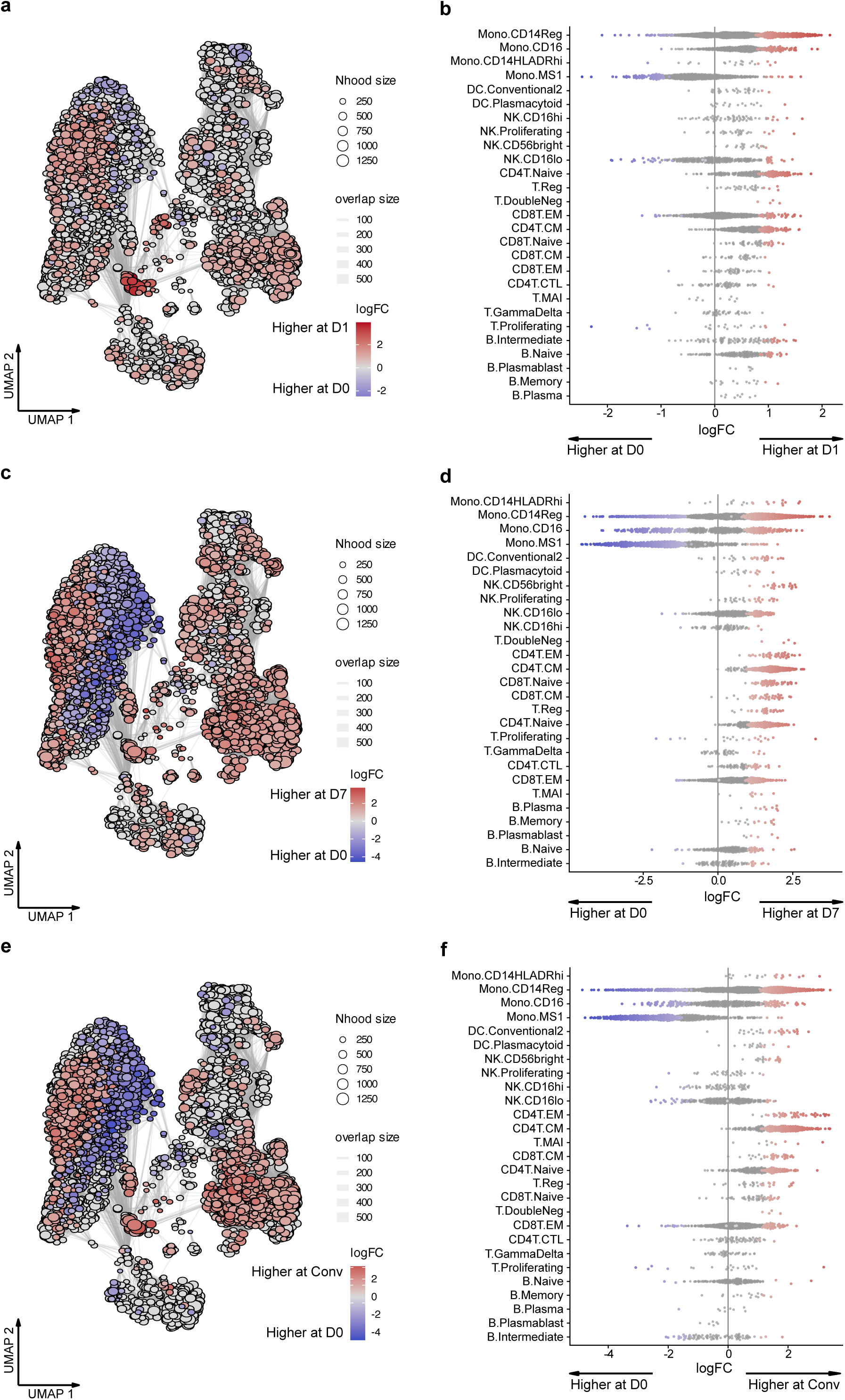
Neighborhood differential abundance across the sepsis time course. Neighborhood graph projection (left of each pair) and beeswarm of per-neighborhood log fold-change grouped by cell lineage (right of each pair) in patients with sepsis, for day 1 versus day 0 (**a**,**b**), day 7 versus day 0 (**c**,**d**), and convalescence versus day 0 (**e**,**f**). Node and point color is the neighborhood log fold-change, with red denoting higher abundance at the later timepoint and blue lower; node size is the neighborhood cell count and edge width the number of cells shared between neighborhoods. Colored beeswarm points are neighborhoods significant at a spatial false discovery rate below 0.1; non-significant neighborhoods are grey. 4,399 neighborhoods were tested in each comparison. D0, D1, and D7 denote days 0, 1, and 7 after presentation; Conv, convalescence; Mono, monocyte; Reg, regular; NK, natural killer; DC, dendritic cell; CTL, cytotoxic T lymphocyte; CM, central memory; EM, effector memory.

**Extended Data Figure 3.**
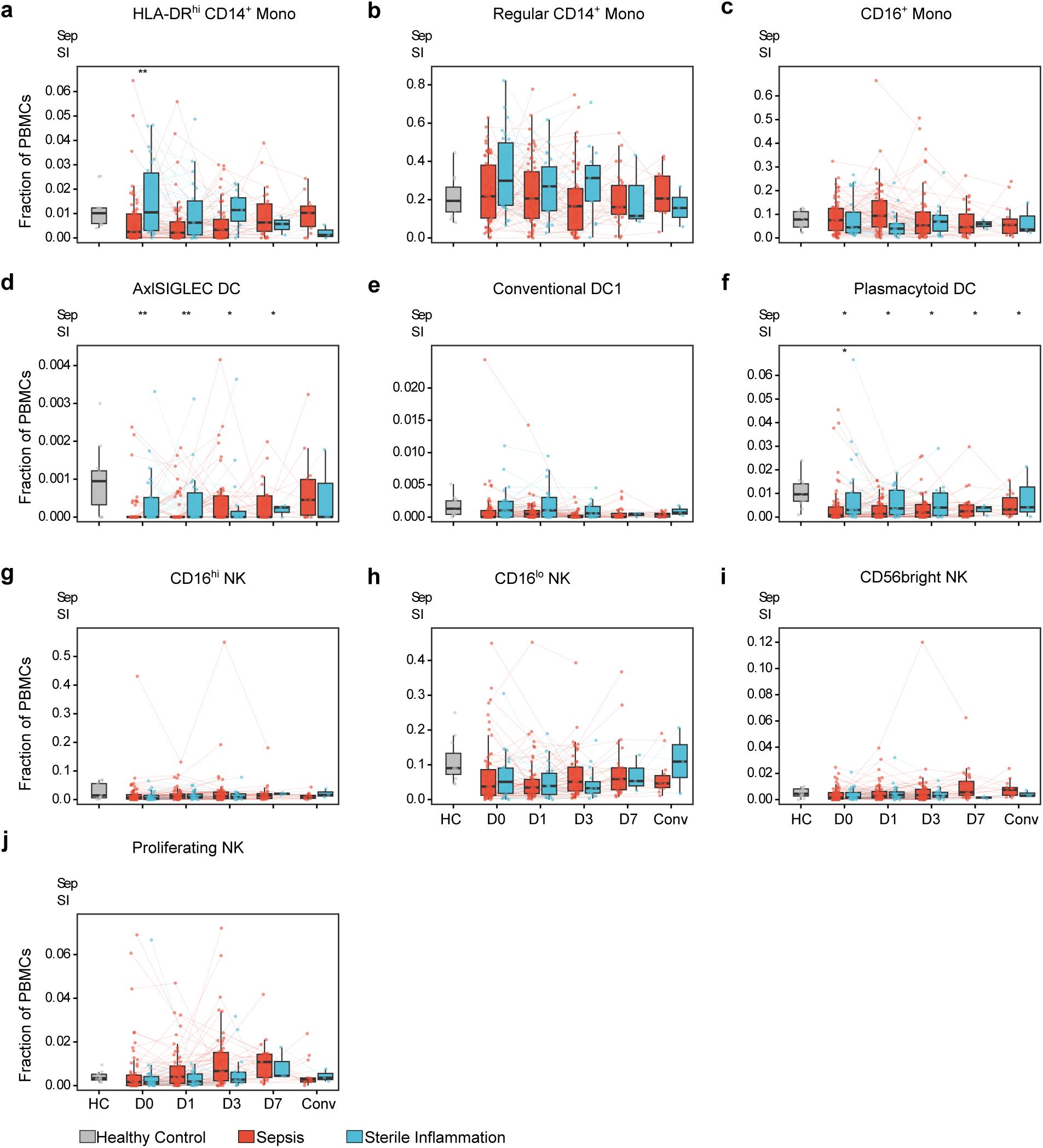
Relative abundance kinetics of monocyte, dendritic cell, and natural killer cell states. Longitudinal trajectories of the fractional abundance of each annotated cell state in patients with sepsis and with sterile inflammation, with healthy controls shown once as a cross-sectional reference. Boxes show median and interquartile range, whiskers extend to 1.5× the interquartile range and thin lines connect samples from the same patient. The upper symbol rows denote significance versus healthy controls for sepsis (“Sep”) and sterile inflammation (“SI”) at each timepoint, and in-plot symbols denote sepsis versus sterile inflammation; comparisons used linear mixed-effects models with a per-patient random effect adjusted for age and sex (* denotes *P_adj_*<0.05, ** *P_adj_*<0.01, *** *P_adj_*<0.001). n = 305 samples from 71 patients with sepsis, 27 patients with sterile inflammation and 12 healthy controls; samples per group at each timepoint are shown in Fig. 1a. D0, D1, D3 and D7 denote days 0, 1, 3 and 7 after presentation; Conv, convalescence; HC, healthy control; SI, sterile inflammation; Mono, monocyte; Reg, regular; NK, natural killer; DC, dendritic cell.

**Extended Data Figure 4.**
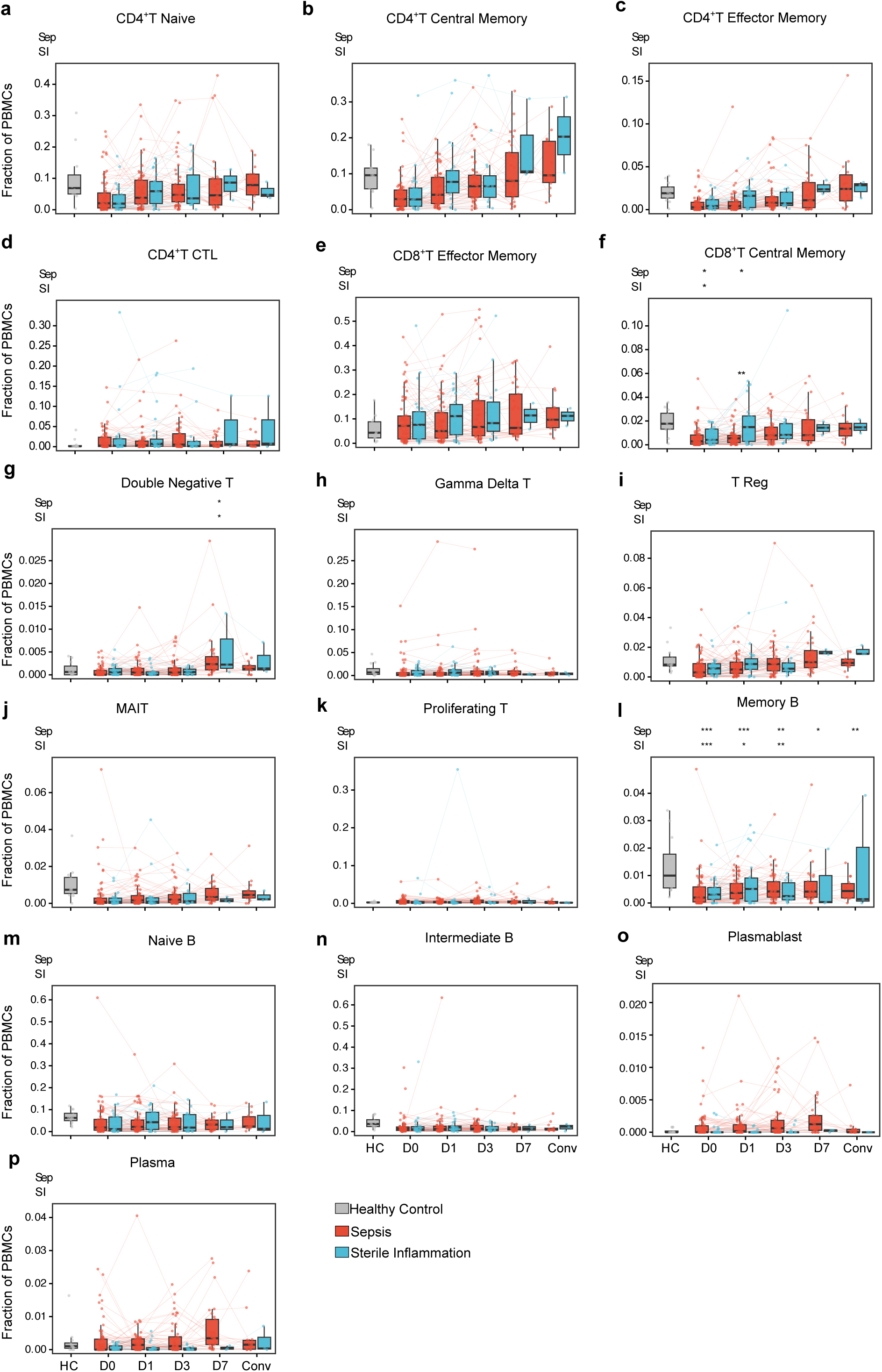
Relative abundance kinetics of T and B cell states. Longitudinal trajectories of the fractional abundance of each annotated cell state in patients with sepsis and with sterile inflammation, with healthy controls shown once as a cross-sectional reference. Boxes show median and interquartile range, whiskers extend to 1.5× the interquartile range and thin lines connect samples from the same patient. The upper symbol rows denote significance versus healthy controls for sepsis (“Sep”) and sterile inflammation (“SI”) at each timepoint, and in-plot symbols denote sepsis versus sterile inflammation; comparisons used linear mixed-effects models with a per-patient random effect adjusted for age and sex (* denotes *P_adj_*<0.05, ** *P_adj_*<0.01, *** *P_adj_*<0.001). n = 305 samples from 71 patients with sepsis, 27 patients with sterile inflammation and 12 healthy controls; samples per group at each timepoint are shown in Fig. 1a. D0, D1, D3 and D7 denote days 0, 1, 3 and 7 after presentation; Conv, convalescence; HC, healthy control; SI, sterile inflammation; CTL, cytotoxic T lymphocyte; MAIT, mucosal-associated invariant T cell.

**Extended Data Figure 5.**
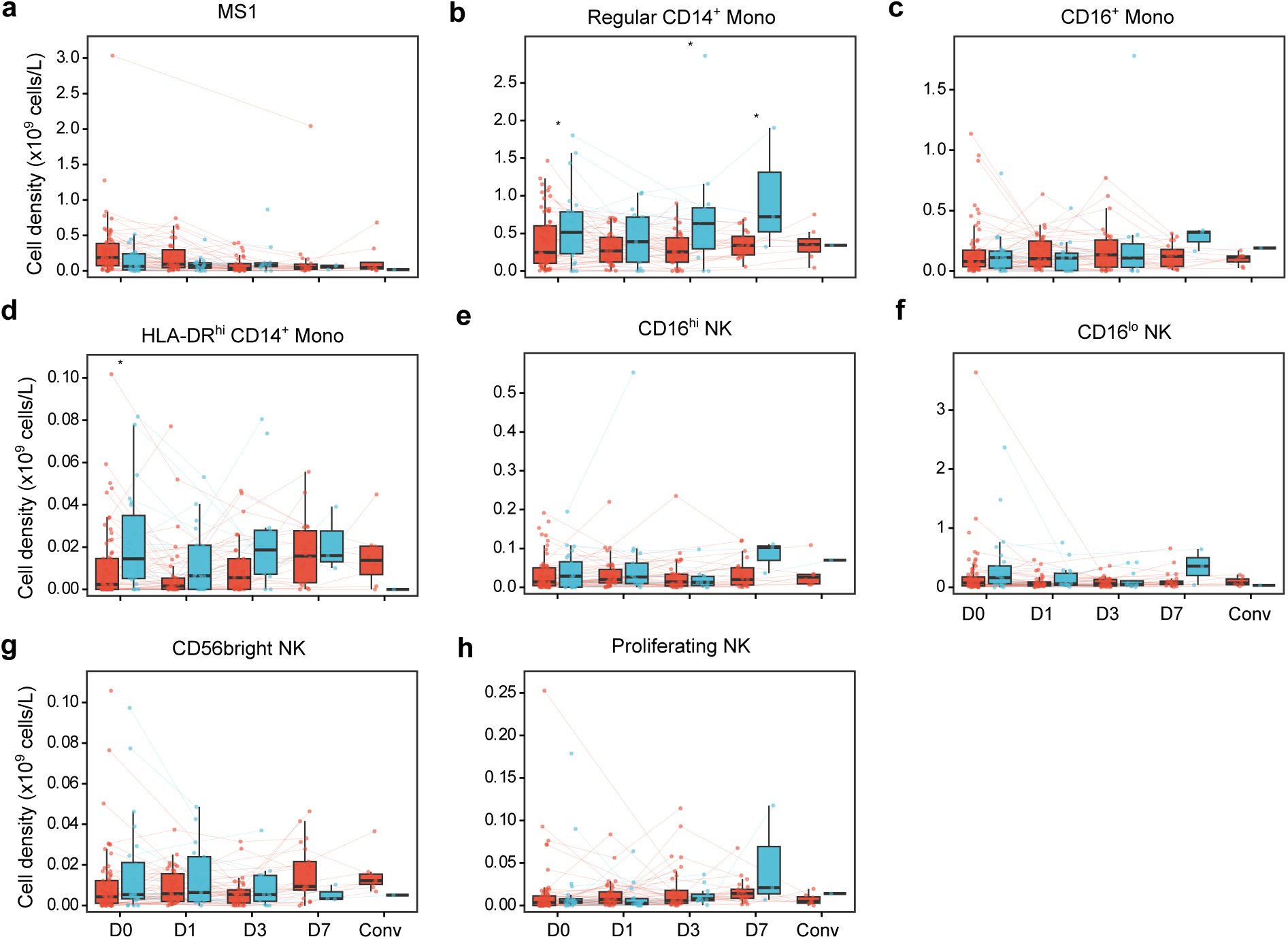
Absolute abundance kinetics of monocyte and natural killer cell states inferred from matched clinical blood counts. Longitudinal trajectories of the inferred absolute abundance (Methods) of each annotated cell state in patients with sepsis and with sterile inflammation. Absolute abundances are not available for healthy controls, who did not have clinical blood counts, or for dendritic cell states, because the clinical differential provides no dendritic cell denominator. Boxes show median and interquartile range, whiskers extend to 1.5× the interquartile range and thin lines connect samples from the same patient. Comparisons used linear mixed-effects models with a per-patient random effect adjusted for age and sex. Samples are those from patients with sepsis or sterile inflammation with a matched clinical blood count. D0, D1, D3 and D7 denote days 0, 1, 3 and 7 after presentation; Conv, convalescence; SI, sterile inflammation; Mono, monocyte; Reg, regular; NK, natural killer.

**Extended Data Figure 6.**
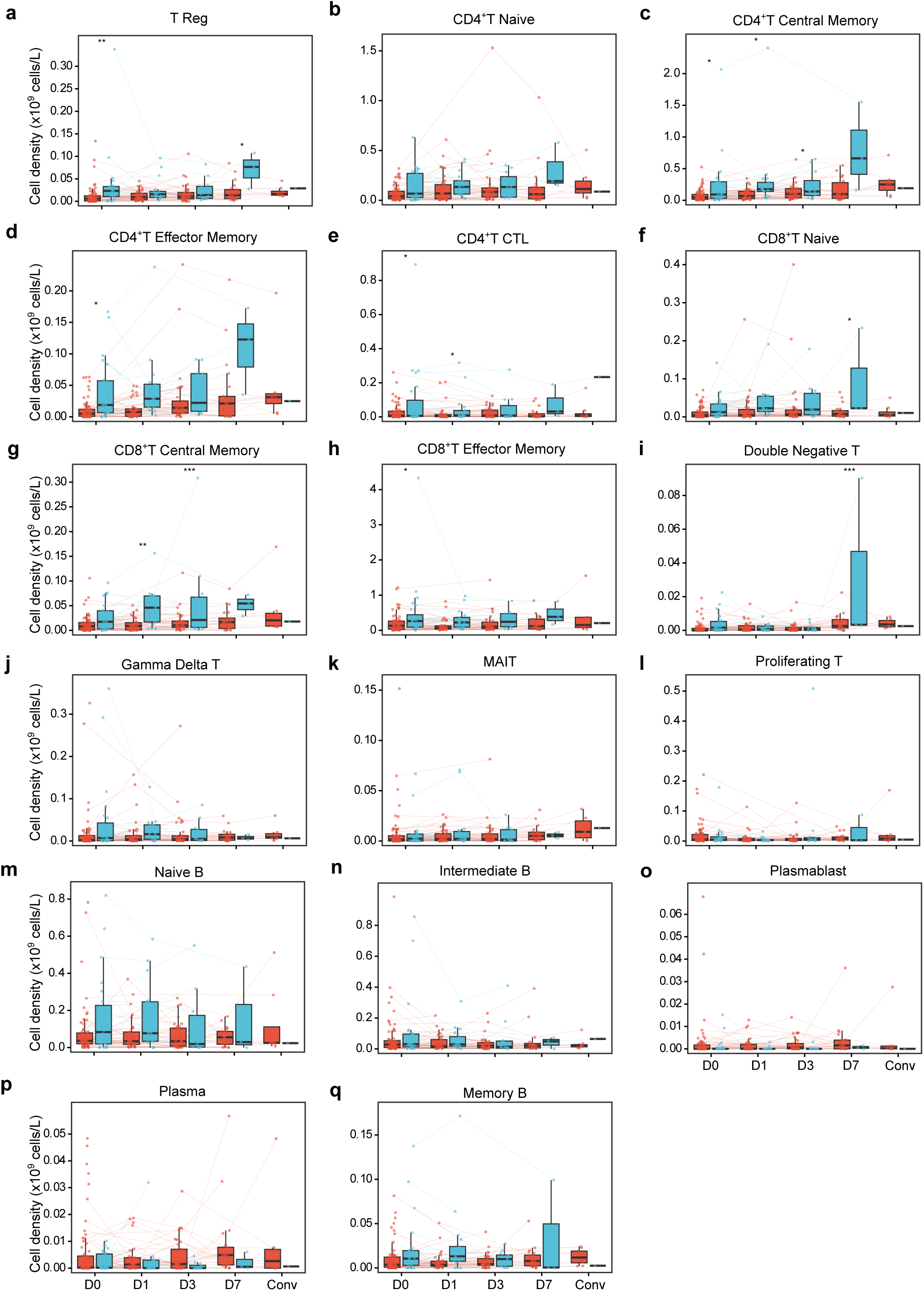
Absolute abundance kinetics of T and B cell states inferred from matched clinical blood counts. Longitudinal trajectories of the inferred absolute abundance (Methods) of each annotated cell state in patients with sepsis and with sterile inflammation. Absolute abundances are not available for healthy controls, who did not have clinical blood counts, or for dendritic cell states, because the clinical differential provides no dendritic cell denominator. Boxes show median and interquartile range, whiskers extend to 1.5× the interquartile range and thin lines connect samples from the same patient. Comparisons used linear mixed-effects models with a per-patient random effect adjusted for age and sex (* denotes *P_adj_*<0.05, ** *P_adj_*<0.01, *** *P_adj_*<0.001). Samples are those from patients with sepsis or sterile inflammation with a matched clinical blood count. D0, D1, D3 and D7 denote days 0, 1, 3 and 7 after presentation; Conv, convalescence; SI, sterile inflammation; MAIT, mucosal-associated invariant T cell; CTL, cytotoxic T lymphocyte.

**Extended Data Figure 7.**
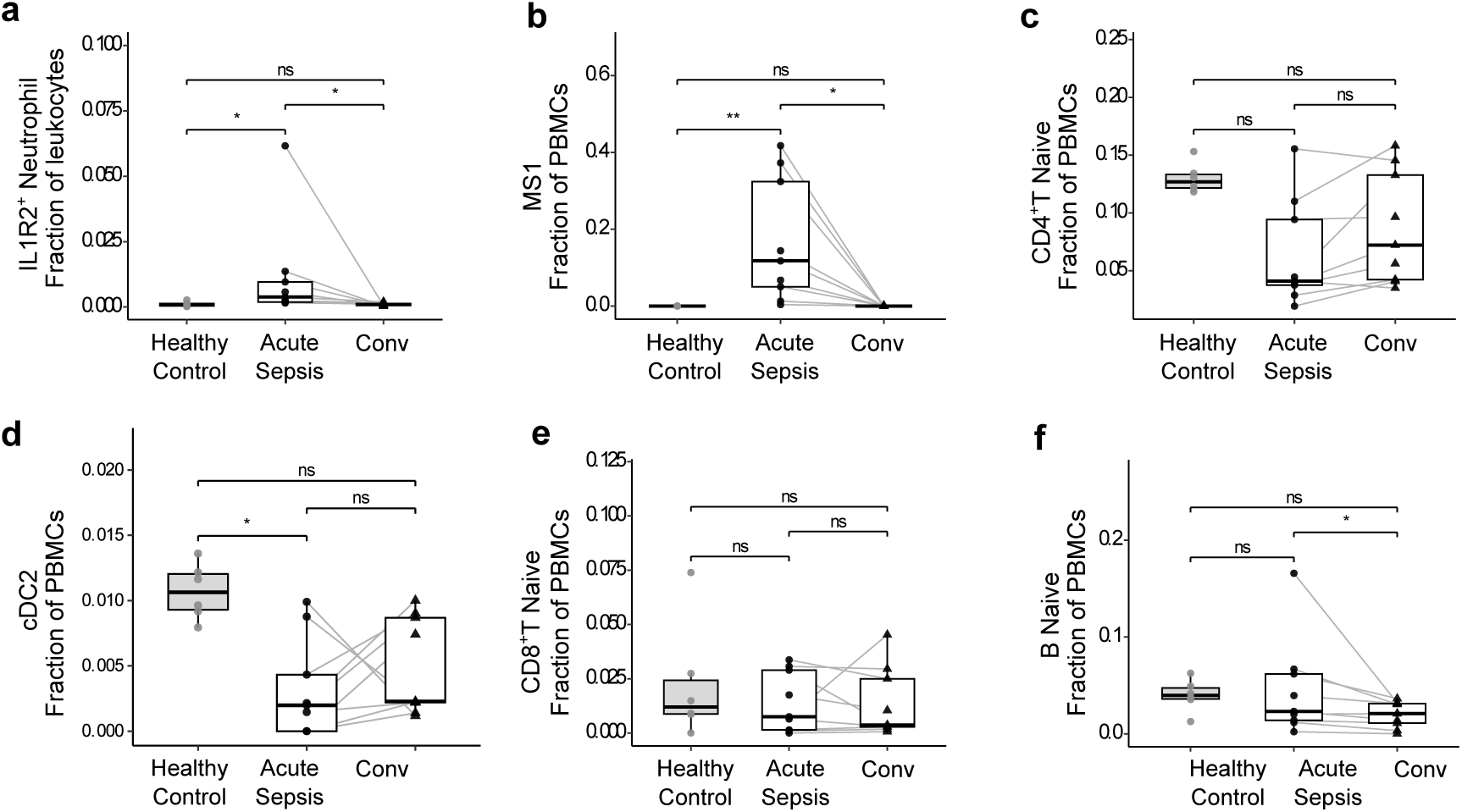
Acute and convalescent abundance of MS1 monocytes, IL1R2⁺ immature neutrophils, and adaptive cell states in an independent sepsis cohort. Reanalysis of the whole-blood single-cell sepsis cohort of Kwok et al.^21^, in which CD14^+^ monocytes were reclustered to resolve an MS1 state. IL1R2^+^ immature neutrophils and adaptive cell states are as annotated in that study. Each panel shows the sample-level abundance of a cell state in healthy controls, acute sepsis and convalescence for: IL1R2⁺ immature neutrophils (**a**), MS1 monocytes (**b**), naive CD4⁺ T cells (**c**), type 2 conventional dendritic cells (**d**), naive CD8⁺ T cells (**e**) and naive B cells (**f**). Brackets show the three pairwise comparisons separately, each a two-sided test with Benjamini–Hochberg adjustment across cell states; * denotes P_adj_ < 0.05, **, *P_adj_* < 0.01, and ns, not significant. n = 9 patients contributed paired acute and convalescent samples. Conv denotes convalescence; MS1, monocyte state 1.

**Extended Data Figure 8.**
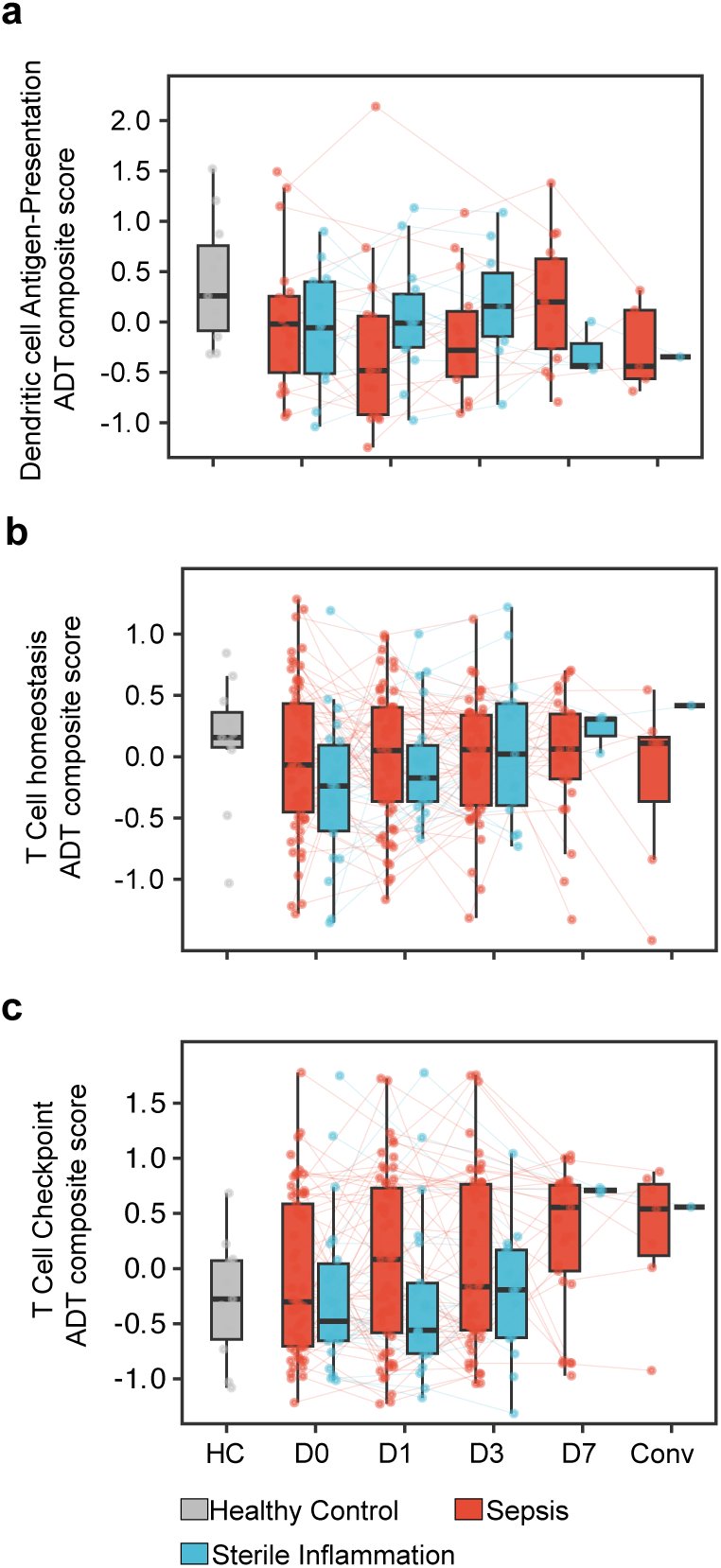
Cell surface protein trajectories. Longitudinal trajectories of composite scores derived from CITE-seq antibody-derived tags in patients with sepsis and with sterile inflammation, with healthy controls shown once as a pooled cross-sectional reference. **a**, dendritic cell (DC) antigen-presentation composite, scored across DC states; **b**, T cell homeostasis and costimulation composite, scored across T cell states; **c**, T cell checkpoint composite, scored across T cell states. Each composite is the mean of z-scored, biologically related surface markers within the indicated cell states (Methods). Boxes show median, interquartile range and 1.5× interquartile range; thin lines connect samples from the same patient. No comparison with healthy controls, or between sepsis and sterile inflammation, was significant at any timepoint (linear mixed-effects models with a per-patient random effect adjusted for age and sex). D0, D1, D3 and D7 denote days 0, 1, 3 and 7 after presentation; Conv, convalescence; HC, healthy control; SI, sterile inflammation; CITE-seq, cellular indexing of transcriptomes and epitopes by sequencing.

**Extended Data Figure 9.**
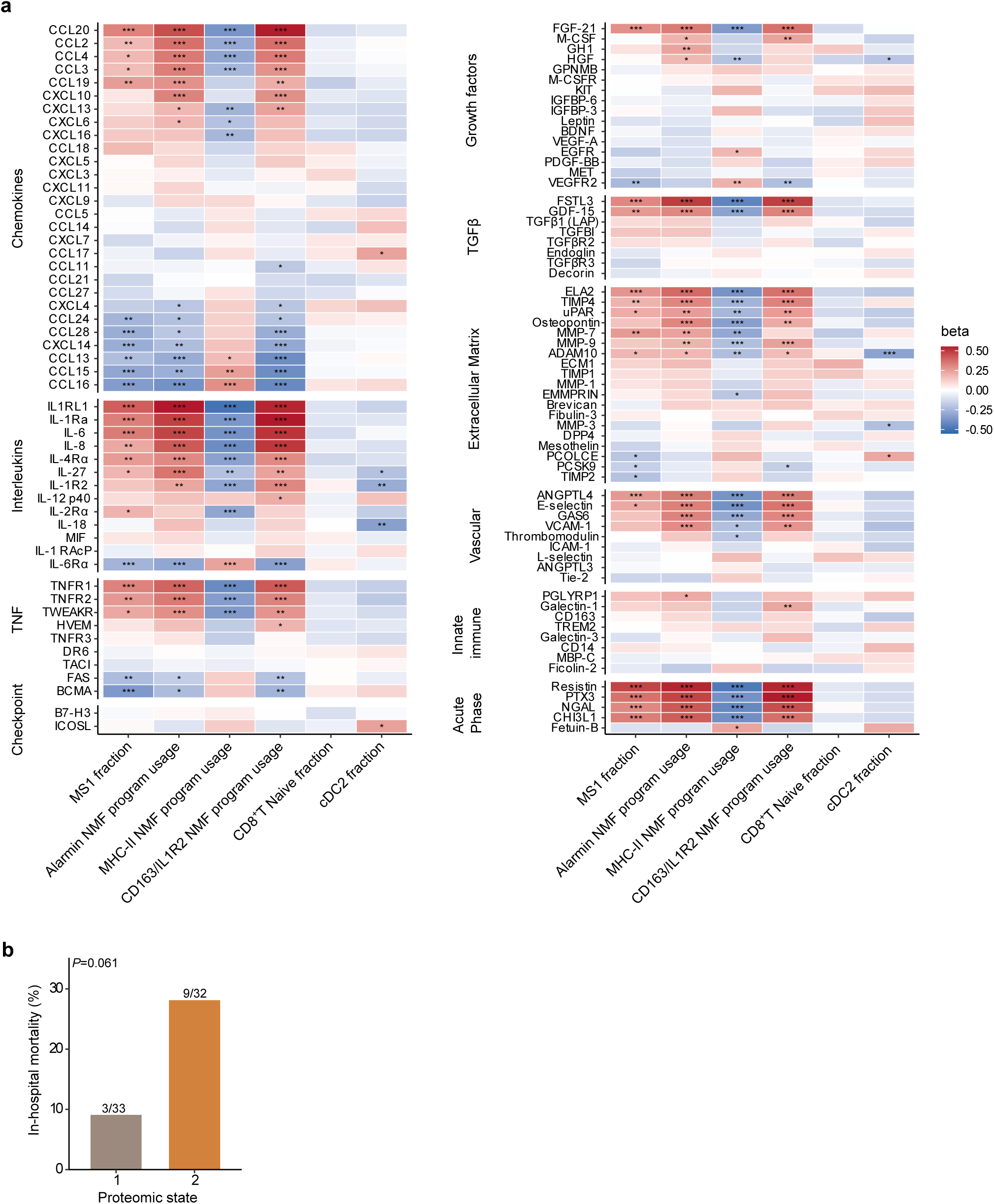
Plasma protein associations with mononuclear features, and in-hospital mortality by plasma proteomic state. **a**, Associations between plasma analytes and PBMC features, estimated across sepsis samples from all timepoints with a per-patient random intercept, adjusted for age and sex. Color shows the standardized coefficient (beta); * denotes a Benjamini–Hochberg false discovery rate (FDR)<0.05, ** FDR<0.01 and *** FDR<0.001. Proteins are grouped by functional class; TNF denotes TNF superfamily proteins; TGFβ, TGFβ and BMP family. **b**, In-hospital mortality in the two plasma proteomic states, with the number of deaths over the number of patients annotated (Fisher’s exact test). n = 65 day 0 sepsis samples with matched plasma, 32 in state 2 and 33 in state 1.

**Extended Data Figure 10.**
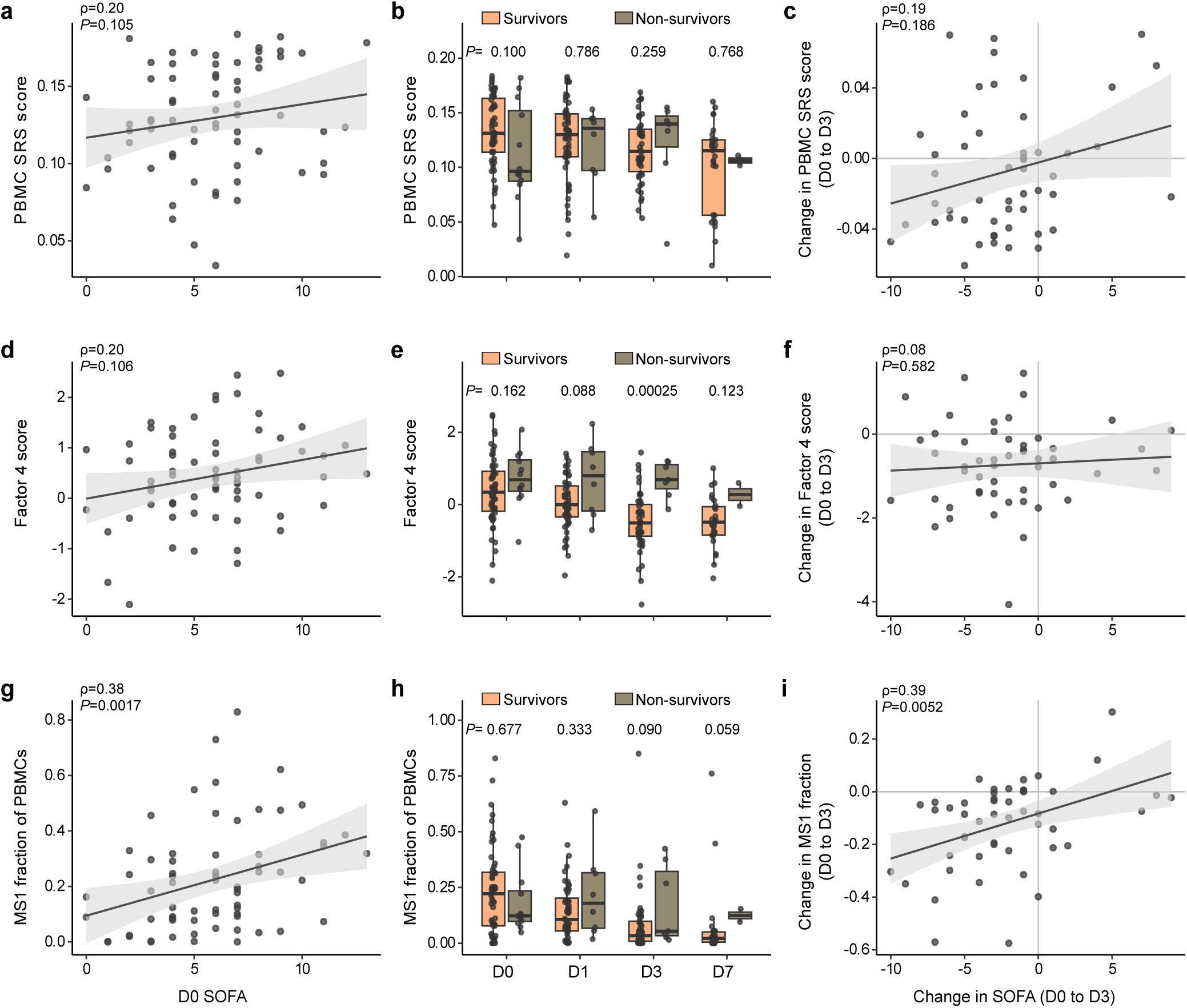
Associations of the PBMC SRS score, the latent host response factor and MS1 abundance with illness severity and in-hospital mortality. Rows are the peripheral blood mononuclear cell (PBMC) sepsis response signature (SRS) score (**a**–**c**), the latent host response factor (Factor 4) (**d**–**f**), and the MS1 fraction of PBMCs (**g**–**i**), in patients with sepsis. **a**,**d**,**g**, Correlation of each measure at presentation with the day 0 Sequential Organ Failure Assessment (SOFA) score; the line is a linear fit with its 95% confidence band and the Spearman correlation coefficient ρ and its *P* value are annotated. **b**,**e**,**h**, Each measure in survivors and in patients who died in the hospital, at days 0, 1, 3 and 7; boxes show median and interquartile range, points are individual samples, and the annotation above each pair is the two-sided Wilcoxon rank-sum *P* value. Panel **e** reproduces Fig. 5e, shown here for direct comparison with the other two measures on identical axes. n = 55, 52, 46 and 27 survivors and 12, 8, 7 and 2 non-survivors at D0, D1, D3 and D7, respectively. Convalescent samples are omitted because only survivors could return for convalescent sampling, leaving no non-survivors at that timepoint. **c**,**f**,**i**, Spearman correlation of the within-patient change in each measure between presentation and day 3 with the parallel change in SOFA score, with ρ and the *P* value annotated. D0, D1, D3 and D7 denote days 0, 1, 3 and 7 after presentation; MS1, monocyte state 1.

## SUPPLEMENTARY FIGURES

**Supplementary Figure 1.**
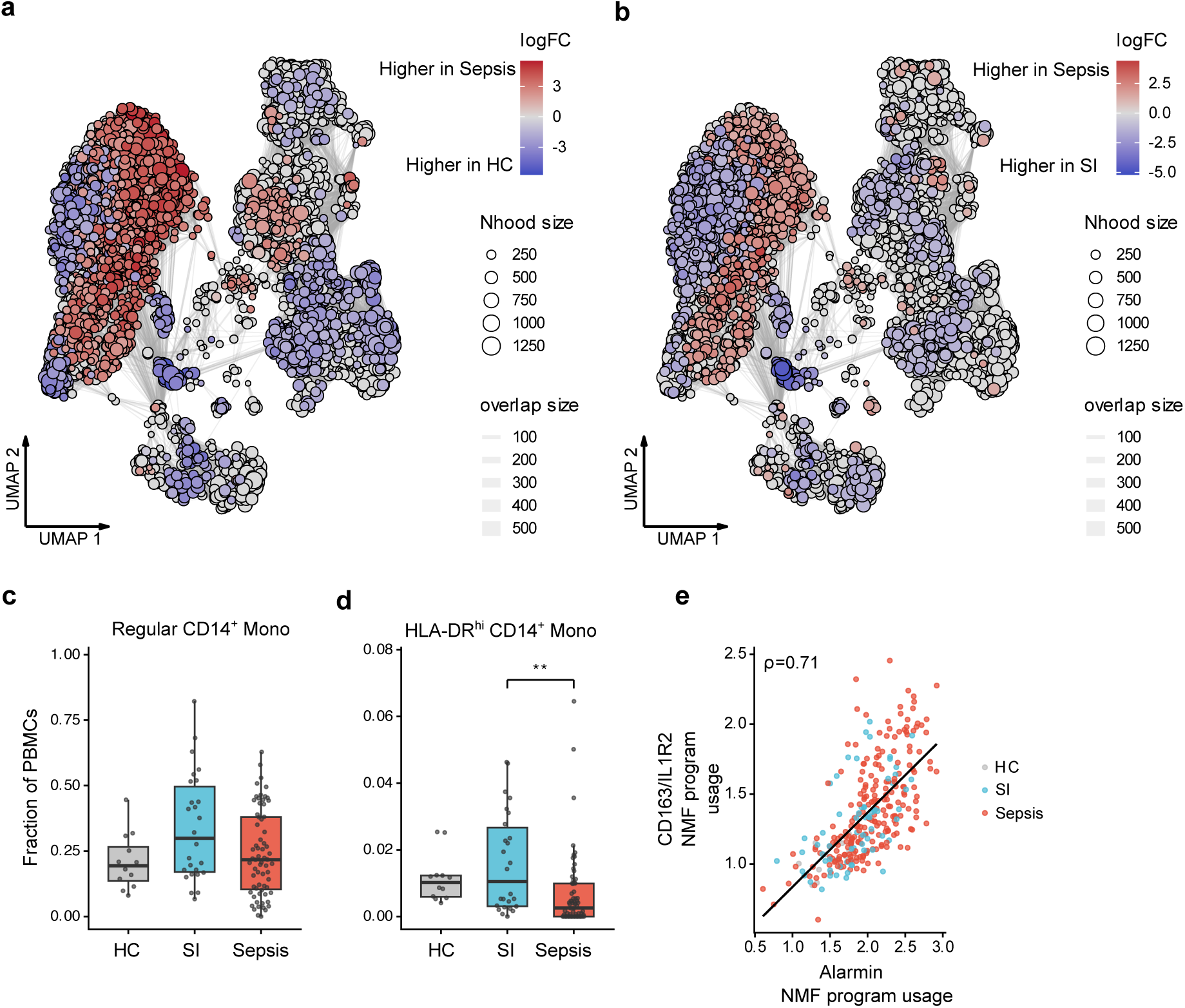
Monocyte remodeling at presentation. **a**,**b**, Neighborhood graph projections of differential abundance at day 0 for sepsis versus healthy controls (**a**) and sepsis versus sterile inflammation (**b**); node color is the neighborhood log fold-change, with red denoting higher abundance in sepsis and blue lower abundance, node size is the neighborhood cell count, and edge width the number of cells shared between neighborhoods. Cell-state labels for the same embedding are in Fig. 1b. **c**,**d**, Regular CD14⁺ (**c**) and HLA-DRʰⁱ CD14⁺ (**d**) monocyte fractions by group at day 0. **e**, Sample-level correlation between alarmin and CD163/IL1R2 program activity in monocytes, colored by group, with Spearman correlation coefficient ρ shown. In **c** and **d**, boxes show median and interquartile range and whiskers extend to 1.5× the interquartile range; in **c**-**e**, each point is one sample. Group comparisons use linear models adjusted for age and sex with Tukey-adjusted contrasts; * denotes *P_adj_* < 0.05, **, *P_adj_* < 0.01, ***, *P_adj_* < 0.001. HC denotes healthy control; SI, sterile inflammation; PBMC, peripheral blood mononuclear cell; Mono, monocyte.

**Supplementary Figure 2.**
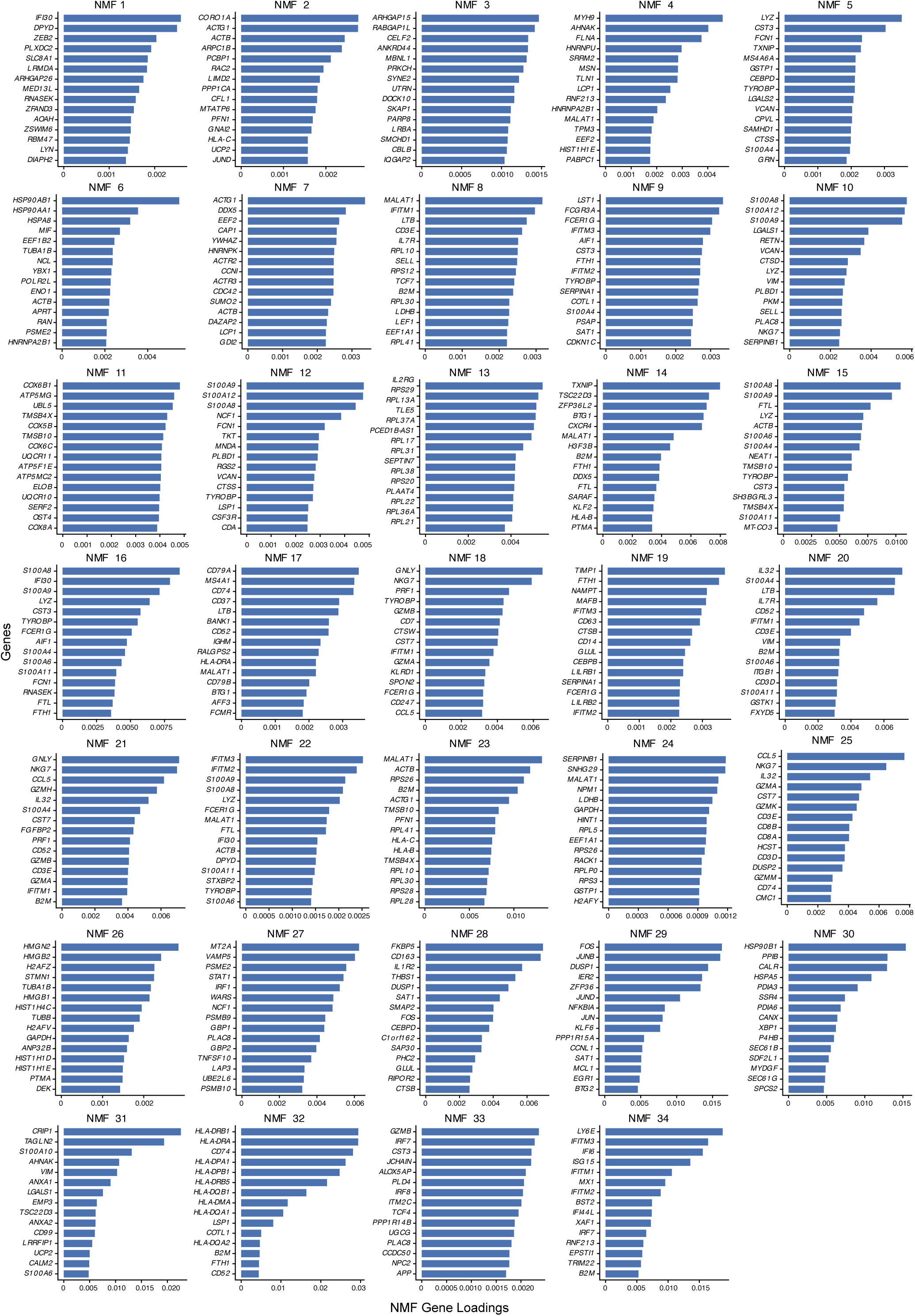
Gene loadings of the non-negative matrix factorization gene programs. Top 15 gene loadings for each gene program derived by non-negative matrix factorization of single-cell transcriptomes from all groups and timepoints. Programs dominated by mitochondrial, ribosomal or hemoglobin genes were considered technical artifacts, were excluded from analysis and are not shown.

**Supplementary Figure 3.**
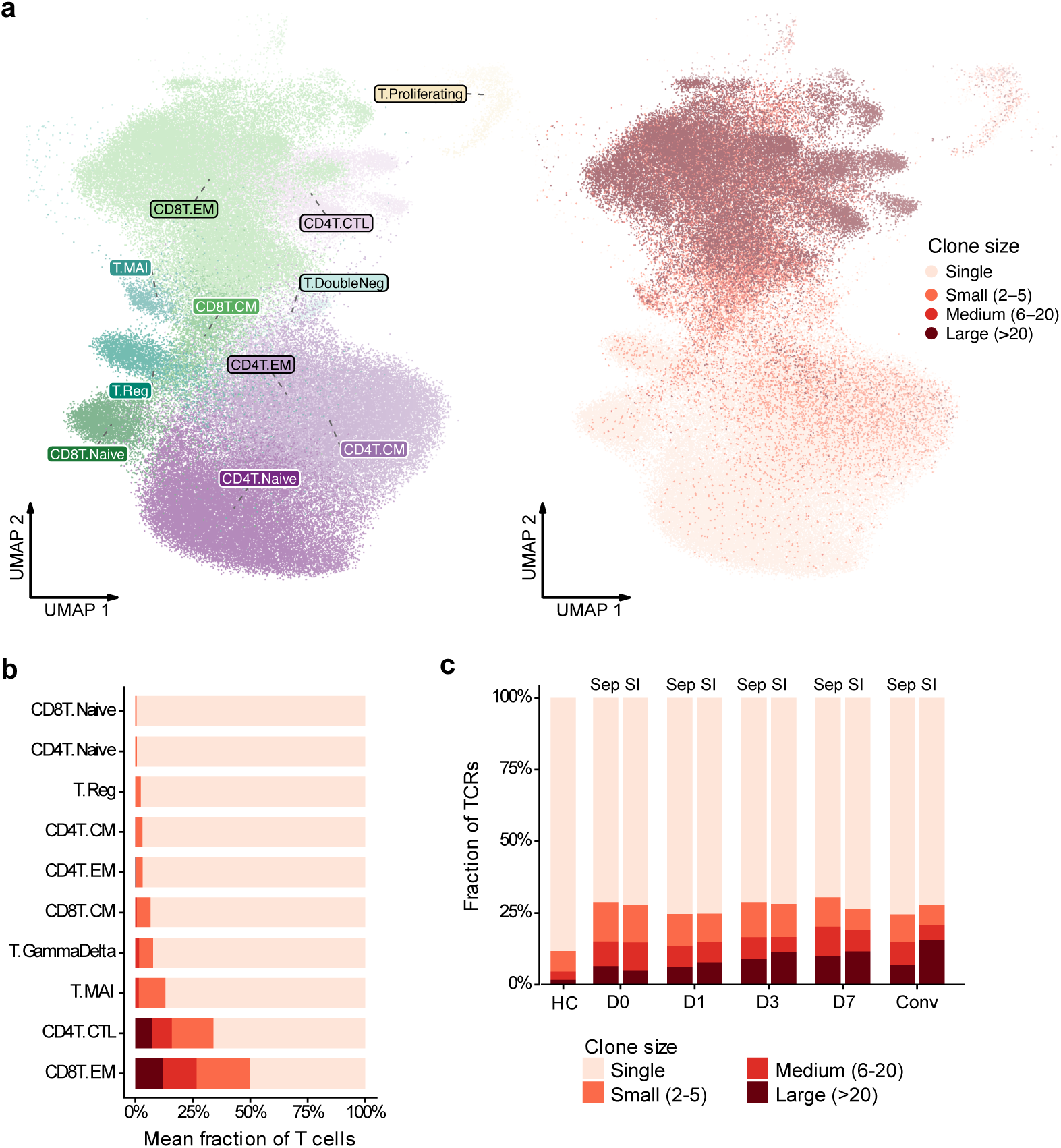
Clonal expansion across T cell states. **a**, Projection of T cells with recovered paired T cell receptor (TCR) sequences, colored by T cell state (left) and by clonotype expansion category (right). **b**, Clone-size composition of each T cell state, as the mean proportion of cells in singleton, small (2–5 cells), medium (6–20 cells) and large (>20 cells) clonotypes; states are ordered by singleton fraction. **c**, Clone-size composition of the total T cell repertoire by group and timepoint. Clonotypes were defined by identical CDR3 nucleotide sequence and V(D)J gene usage across both α and β chains. D0, D1, D3 and D7 denote days 0, 1, 3 and 7 after presentation; Conv, convalescence; HC, healthy control; SI, sterile inflammation; Sep, sepsis; CM, central memory; EM, effector memory; CTL, cytotoxic T lymphocyte.

**Supplementary Figure 4.**
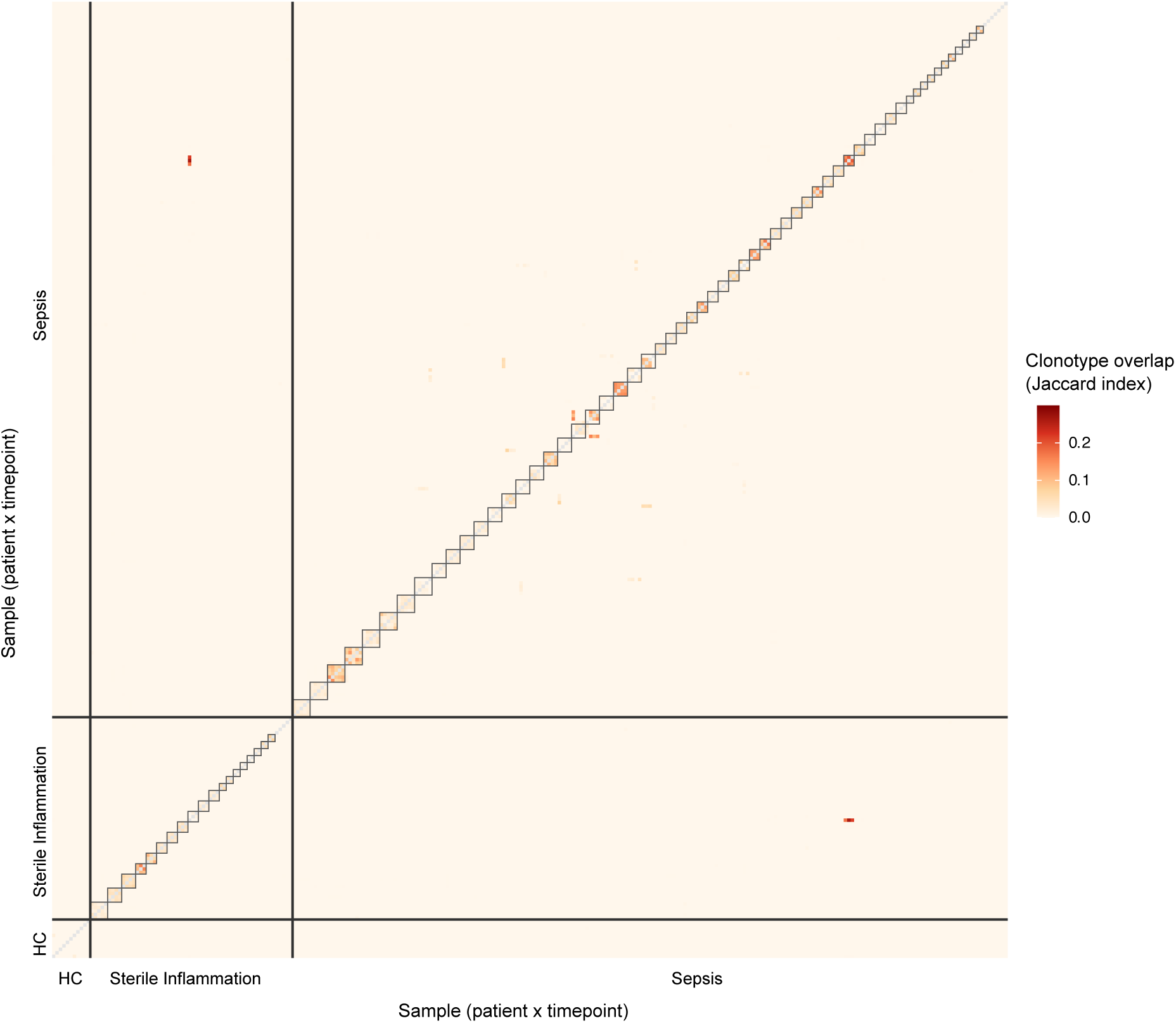
T cell receptor repertoire overlap between patients. Pairwise Jaccard overlap of clonotype sets between all samples, grouped by healthy control (HC), sterile inflammation, and sepsis; boxes on the diagonal demarcate samples from the same patient.

**Supplementary Figure 5.**
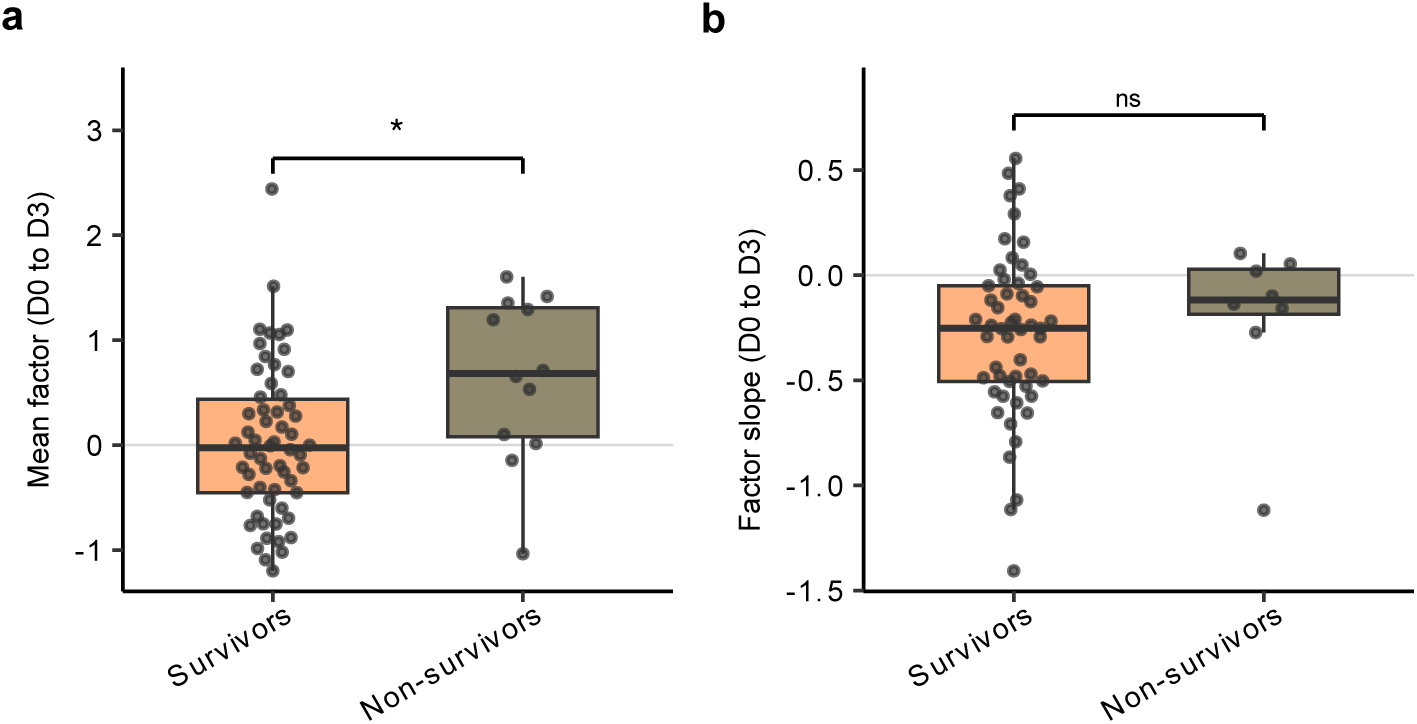
Latent host response factor (Factor 4) scores between presentation and day 3, by outcome. **a**, Mean factor value across days 0 to 3 in sepsis survivors and in patients who died in the hospital (n = 58 and 12 respectively). **b**, Per-patient rate of change in Factor 4 between day 0 and day 3, estimated by linear regression on each patient’s samples, in survivors and non-survivors. Boxes show median and interquartile range, whiskers extend to 1.5× the interquartile range and points are individual patients. Groups were compared by two-sided Wilcoxon rank-sum tests (* denotes P<0.05, ns, not significant). D0 and D3 denote days 0 and 3 after presentation.

## SUPPLEMENTARY TABLES

**Supplementary Table 1. Leave-one-healthy-control-out sensitivity analysis**. Each healthy-control-referenced contrast re-estimated with one control omitted at a time, reporting the full-model standardized estimate and Benjamini–Hochberg-adjusted *P* value alongside the fraction of iterations preserving the sign and the significance of the effect, and the median and 5th to 95th percentile range of the leave-one-out estimates. n = 12 healthy controls, giving 12 iterations per contrast.

**Supplementary Table 2. T cell receptor sequencing depth and repertoire diversity**. Panel A reports the number of cells with a recovered paired T cell receptor per sample by group and timepoint. Panel B lists every group and timepoint contrast for the overall T cell repertoire at Hill orders q = 2 (the primary metric), 1 and 0, with and without adjustment for log sequencing depth, and the Spearman correlation between each diversity metric and depth; the depth-adjusted columns represent the primary analysis. Panel C repeats the q = 2 contrasts after rarefying every sample to a common depth of 25 or 50 cells. Panel D, on a second tab, reports for each T cell subset the median number of TCR-bearing cells per sample, the proportion of samples in which every clonotype was observed once and the rank correlation between the q = 2 estimate and the number of cells sampled, together with the subset-level q = 2 contrasts. *P* values are Benjamini–Hochberg adjusted across the contrasts within one diversity metric and estimator.

**Supplementary Table 3. Plasma protein modules**. Proteins assigned to four modules derived by correlation-based clustering, across samples from patients with sepsis, of the plasma analytes passing the prespecified detectability filter.

**Supplementary Table 4. Benchmarking of PBMC-derived SRS-signature recovery against a whole-blood single-cell reference**. Panel A compares discrimination of SRS1 from non-SRS1 samples for the supervised SepstratifieR classifier and for the PBMC sepsis response signature score, each applied to whole blood and to mononuclear pseudobulk derived from the same cells, giving the area under the receiver operating characteristic curve, accuracy, sensitivity and specificity. Panel B shows the correlation of each score with the reference SRS score (SRSq). n = 26 samples (16 SRS1, 10 non-SRS1) from the reference cohort. PBMC denotes peripheral blood mononuclear cell; SRS, sepsis response signature.

**Supplementary Table 5. All latent factors of the multi-omics factor analysis model.** For each of the eight factors of the model fitted on samples from patients with sepsis only: the sum of signed Spearman correlations with six prespecified features (selection score), the percentage of variance explained in each group of inputs, and its association with sampling day (Spearman), with SOFA score and the PBMC SRS score (linear mixed-effects models adjusted for sampling day, per-patient random intercept) and with in-hospital mortality (across timepoints by mixed-effects model, and at each timepoint by two-sided Wilcoxon rank-sum test). *P* values are shown unadjusted and Benjamini–Hochberg adjusted across the eight factors. Factors are oriented by their agreement with the prespecified features.

**Supplementary Table 6. Associations of the latent host response factor with all input features**. Every feature that entered the multi-omics factor analysis model, with its data view and its association with the factor, adjusted for sampling day, unadjusted and estimated within patients, with Benjamini–Hochberg-adjusted *P* values.

## METHODS

### Study Ethics, Patient Cohorts, and Sample Collection

Adults (≥18 years of age) presenting to the Emergency Department at Massachusetts General Hospital (Boston, MA) between 1 August 2018 and 30 June 2023 were screened for enrollment. All study procedures were conducted in accordance with the Declaration of Helsinki and were approved by the Institutional Review Boards of the Broad Institute of MIT and Harvard and of Massachusetts General Hospital. Written informed consent was obtained from all participants or their legally authorized surrogates at enrollment or, when necessary, retrospectively after recovery of decision-making capacity. Participants were adjudicated into one of two primary clinical categories: sterile inflammation or sepsis. Adjudication was performed independently by three physician-investigators blinded to the molecular data; the final classification was based on microbiological and laboratory findings, response to therapy and clinical evolution during hospitalization, with discrepancies resolved by consensus. The sepsis group comprised patients meeting Sepsis-3 criteria^43^, and the sterile inflammation group comprised patients with evidence of systemic inflammation without infection. To minimize treatment-induced confounding, patients were excluded if they had received any antimicrobial within 7 days before presentation, or intravenous antibiotics for more than 12 hours while in the ED before enrollment. Illness severity was quantified by the Sequential Organ Failure Assessment (SOFA) score, calculated daily from clinical and laboratory data. In-hospital mortality was assessed via chart review. Sex was obtained from the electronic health record. The sepsis group comprised 27 female and 44 male patients, the sterile inflammation group 11 female and 16 male patients, and the healthy control group 6 female and 6 male participants. Sex was included as a covariate in all cross-sectional and longitudinal models; the study was not designed or powered to test for sex differences in the host response, and no sex-disaggregated analyses were performed. Race and ethnicity were self-reported by participants from fixed categories; they were not used in any model.

Peripheral blood was collected at enrollment (day 0), corresponding to initial Emergency Department presentation, and subsequently on days 1, 3, and 7 for hospitalized patients when feasible. Convalescent blood samples were obtained from a subset of participants at least 28 days after enrollment; the exact timing of convalescent sampling varied among participants. Healthy control samples were obtained from Research Blood Components (Watertown, MA), and were age- and sex-matched to disease cohort participants where feasible. Healthy controls were sampled at a single timepoint and processed using the same workflow as clinical samples.

### Blood Processing, Cryopreservation, and Single-cell Library Preparation and Profiling

Peripheral blood was collected by venipuncture into ethylenediaminetetraacetic acid (EDTA) vacutainers (BD Biosciences). Whole blood was diluted 1:1 with phosphate-buffered saline (PBS) and subjected to density gradient centrifugation using Ficoll-Paque Plus (Cytiva), following the manufacturers’ protocols. After centrifugation at 1,200 × g for 20 minutes at room temperature with the brake off, the peripheral blood mononuclear cell (PBMC) layer was collected and washed in RPMI 1640 (Gibco) with 2% fetal bovine serum (FBS). Cells were counted using an automated cell counter and resuspended in CryoStor CS10 (STEMCELL Technologies). Aliquots containing approximately 1 × 10^6^ PBMCs were prepared in cryovials, cooled overnight at −80°C in controlled-rate containers (Corning CoolCell) and transferred to liquid nitrogen for long-term storage. Plasma was collected in parallel from the Ficoll supernatant, aliquoted and stored at −80 °C until analysis.

For single-cell analyses, PBMCs were thawed in a 37°C water bath for 1–2 minutes and washed twice in RPMI + 10% FBS or Fluorescence-Activated Cell Sorting (FACS) buffer (PBS with 2% FBS and 2 mM EDTA). For immunophenotyping and hashing prior to multiplexed single-cell RNA sequencing, cells were stained with a viability dye (4′,6-diamidino-2-phenylindole [DAPI], ThermoFisher) and fluorescent-conjugated antibodies (Alexa Fluor 700 anti-CD15 and FITC anti-CD235a, BioLegend) to allow exclusion of dead or contaminating non-PBMC populations. Fc receptors were blocked with Human TruStain FcX (BioLegend). Hashtag oligonucleotide-conjugated antibodies (TotalSeq™ anti-human Hashtag, BioLegend) were used to uniquely label samples before pooling. Surface proteins were profiled with the TotalSeq-C Human Universal Cocktail v1.0 (BioLegend), a panel of oligonucleotide-conjugated antibodies compatible with 5′ capture comprising 130 antibodies and 7 isotype controls, used according to the manufacturer’s protocol. Sorting was performed on a SONY MA900 or BD FACSAria II cell sorter using a 100-µm nozzle, with collection into chilled RPMI + 10% FBS. Gating excluded debris and doublets based on forward and side scatter, and excluded DAPI-positive dead cells. Sorted cells were counted, washed, and pooled for library preparation.

Single-cell RNA and surface protein profiling were performed using 10x Genomics Chromium Single Cell 5’ v2 chemistry, with parallel generation of gene-expression, hashtag oligonucleotide and antibody-derived tag (ADT) libraries. T cell receptor libraries were prepared from the same amplified cDNA by targeted enrichment of TCR α and β chain transcripts with the Chromium Single Cell Human TCR Amplification Kit (10x Genomics), following the manufacturer’s protocol, and were sequenced to a target depth of approximately 5,000 read pairs per cell. Final libraries were pooled to achieve target depths of approximately 50,000 reads per cell for gene expression and approximately 10,000 reads per cell for ADT and hashtag libraries, and were sequenced on an Illumina NovaSeq S4 flow cell.

### Single-cell preprocessing and cell-state annotation

FASTQ files were processed using Cell Ranger (v6.0) with alignment to the GRCh38 human reference genome. TCR sequences were assembled and annotated with Cell Ranger vdj (v6.0) against the GRCh38 V(D)J reference, and clonotype analyses were restricted to cells with a productive TCR α and β chain that also passed transcriptome quality control. Multiplexed samples were demultiplexed using cell hashing, and doublets were identified using HTODemux in Seurat. Cells with discordant RNA, ADT and hashtag identities or high background signal were removed. RNA data were log-normalized, and ADT counts were normalized using the centered log-ratio transformation. Dimensionality reduction was performed by principal component analysis, and integrated multimodal neighbor graphs were constructed using Seurat weighted nearest-neighbor (WNN) analysis. Batch correction across donor and timepoint used reciprocal principal component analysis. Broad immune cell types and finer subclusters were annotated manually using canonical RNA and ADT markers, supported by projection onto external reference atlases^44^.

### Neighborhood-based Differential Abundance Analysis

To identify differentially abundant immune cell neighborhoods across phenotypes, we used MiloR (v1.3.0)^29^. A shared nearest-neighbor graph was constructed from the WNN reduction that jointly incorporated RNA and ADT modalities, and neighborhoods were defined using graph-based refinement. Neighborhood counts were computed at the sample level. Differential abundance testing used phenotype or timepoint as the variable of interest with age and sex as covariates; timepoint contrasts were constructed as the later timepoint minus day 0, and the log fold-change is reported unmodified, so positive values indicate higher abundance at the later timepoint. *P* values were adjusted using overlap-aware, graph-based weighting, and significant neighborhoods were defined by a spatial false discovery rate (SpatialFDR) below 0.1. Each neighborhood was annotated according to its dominant cell state composition. States contributing fewer than three neighborhoods were omitted from beeswarm displays, and the ordering within lineage uses the signed median log fold-change over significant neighborhoods.

### Non-negative Matrix Factorization and Gene Program Scoring

To identify transcriptional programs underlying immune cell heterogeneity, we performed non-negative matrix factorization (NMF) on log-normalized single-cell RNA-seq data using the singlet package (v0.2.4)^30^. The top-loading genes from each retained factor were used to define gene programs; modules dominated by mitochondrial, ribosomal or hemoglobin genes were considered technical and excluded. To quantify gene program activity at the sample level, log-normalized expression values were averaged across all cells from the same sample within each immune cell lineage to generate sample-level pseudobulk profiles, and program scores were computed as weighted averages of the top-loading genes using normalized NMF gene weights.

### Surface protein composites

Cell-surface protein (ADT) abundance was summarized per sample within each transcriptionally defined cell state (Fig. 1b) as the mean centered log-ratio-normalized value across the cells of that state, so that each state in each sample contributed one observation rather than its constituent cells. To summarize coordinated surface-protein programs, composite scores were defined as a prespecified group of markers together with the lineage whose constituent cell states they were scored in: the T cell homeostasis and costimulation composite comprises IL7R, SELL, CD28 and ICOS, the T cell checkpoint composite PDCD1, TIGIT, LAG3 and CTLA4, both scored across all T cell states; the monocyte antigen-presentation composite HLA-DRA, HLA-A and CD86, scored across all monocyte states; and the dendritic cell antigen-presentation composite CD1C, ITGAX, HLA-DRA and CD86, scored across all dendritic-cell states. The abundance of each marker in each cell state was z-scored across samples, and the composite is the mean of these z-scores over every marker in the group and every cell state in which it was scored, yielding one score per sample for cross-sectional and longitudinal analysis.

### Inference of Absolute Immune-Cell-State Abundance

To estimate absolute counts of immune-cell subsets, we combined clinical complete blood count (CBC) data with single cell state proportions. Absolute monocyte and lymphocyte counts (×10⁹ cells/L) were obtained from matched CBC-with-differential data generated by clinical laboratories at the time of sample collection. For each sample, the inferred absolute count of a monocyte state was calculated as its proportion among all monocytes multiplied by the total absolute monocyte count. An analogous approach was applied to lymphocyte populations: the proportion of each T cell, B cell or natural killer cell state within the transcriptomically defined lymphocyte pool was multiplied by the corresponding total absolute lymphocyte count. Dendritic cell states have no absolute counterpart because the clinical differential provides no dendritic cell denominator. Samples lacking matched complete blood count data were excluded from absolute-count analyses.

### T cell receptor repertoire analysis

T cell receptor (TCR) analyses were performed on paired single-cell transcriptome and TCR data from transcriptionally defined T cell states. Clonotypes were defined by identical CDR3 nucleotide sequence and V(D)J gene usage across both α and β chains. TCR-bearing cells were grouped into clone-size classes (single, small, medium, and large), and clone-size composition was summarized within each T-cell state and at the sample level. To quantify repertoire diversity, we calculated Hill diversity indices at orders q = 0, q = 1 and q = 2 for the overall T cell repertoire and for individual T cell subsets. Hill q = 2 was designated the primary diversity metric before analysis because it weights abundant clonotypes and is therefore sensitive to redistribution of repertoire space by selective clonal expansion, and also because it is the least sensitive member of the Hill family to incomplete sampling^31^. Per-sample TCR-bearing cell counts are reported, and all diversity contrasts include log-transformed depth as a covariate. As a sensitivity analysis, the q = 2 contrasts were repeated after rarefying every sample to a common depth of 25 or 50 TCR-bearing cells. Pairwise clonotype sharing between samples was quantified using the Jaccard index.

### Inference of a PBMC-compatible sepsis response signature (SRS) surrogate score

To determine a PBMC-compatible surrogate for the whole-blood SRS endotype signature, we used a reference whole blood single cell dataset with sample-level SRS labels and a quantitative SRS score (SRSq)^21,33^. For each reference sample, pseudobulk expression profiles were generated from single-cell RNA, and candidate PBMC surrogates were evaluated against the reference SRS1 versus non-SRS1 classification and SRSq using receiver operating characteristic area under the curve and Pearson and Spearman correlations across the reference samples. We compared whole-blood and PBMC-restricted implementations of the SepstratifieR SRS classification algorithm^33^ together with a PBMC pseudobulk surrogate based on the extended 19-gene SRS signature^33^. The 19-gene PBMC pseudobulk surrogate showed the strongest transferable performance and was therefore selected for downstream analyses.

### Plasma Proteomics

Plasma samples were profiled using the Nucleic Acid-Encoded Library Immunoassay (nELISA), a high-throughput bead-based platform for multiplexed quantification of protein targets, which uses DNA-barcoded antibody pairs and DNA strand-displacement chemistry with readout by flow cytometry^45^. Sample processing, hybridization, and decoding were performed according to manufacturer protocols (Nomic Bio, Montreal, Canada). Analyte panels included pro- and anti-inflammatory cytokines, growth factors, chemokines, and tissue remodeling proteins. Raw intensity values were normalized using internal controls and standard curves to generate log-transformed concentrations. For analytes with concentrations falling outside the assay’s dynamic range, values below the lower limit of detection (LLOD) were imputed as the LLOD, and values above the upper limit of detection (ULOD) were imputed as the ULOD, following the manufacturer’s guidelines. All downstream analyses used log10-transformed concentrations unless otherwise specified. Because analytes quantified near the limits of detection contribute disproportionate technical noise, and many inflammatory mediators are only transiently elevated in the early acute phase of sepsis^46^, we applied a prespecified detectability filter that excluded proteins with more than 30% of day 0 values outside the assay’s lower or upper limits of detection, retaining proteins reliably quantified in at least 70% of presentation samples (117 of 264 measured proteins). This threshold is consistent with detectability-based filters applied in comparable plasma-proteomic studies of sepsis and critical illness^47^ and with the limits-of-detection framework of the nELISA platform^45^.

Associations between plasma proteins and transcriptomic features were estimated within sepsis samples from all timepoints, on log₁₀-transformed concentrations of the 117 proteins passing the detectability filter, with a per-patient random intercept and adjustment for age and sex. P values were adjusted by the Benjamini–Hochberg method across proteins within each feature. Proteomic states were derived from the 117 proteins passing the detectability filter, by k-means clustering of day 0 sepsis samples on the first 14 principal components, which together explained 75% of the variance, with k = 2 chosen by silhouette width; all remaining samples, including healthy controls and sterile inflammation, were projected into the same space and assigned to the nearer centroid without contributing to the partition. Plasma protein modules were defined by hierarchical clustering of proteins on Spearman correlation across longitudinal sepsis samples, sample-level module scores were calculated as the mean z-scored abundance of proteins within each module, and associations between module scores and PBMC features were tested using mixed-effects models.

### Multi-omics factor analysis

We performed targeted multi-omic latent-factor analysis using MOFA2^34^ on sample-level data restricted to sepsis samples. The model was fitted on all 223 sepsis samples, with missing values modeled rather than excluded, across five views and 39 features, using a fixed random seed and a maximum of 2,000 training iterations. The views were PBMC cell state abundances, NMF program scores, TCR diversity metrics, ADT composites, and plasma proteomic features. Preliminary fits across a range of factor numbers showed that, with eight factors, each factor explained at least 5% of the variance of at least one view; the number of factors was therefore fixed at eight, and default MOFA2 settings were applied for all other parameters. Each of the eight factors was scored against six features of the response specified before the model was fitted: higher MS1 abundance and higher monocyte alarmin program activity, together with lower monocyte MHC class II program activity and lower abundance of CD4⁺ central memory, CD4⁺ effector memory and CD8⁺ central memory T cells. Each factor was first oriented so that its overall agreement with this set was positive, then scored as the sum of its Spearman correlations with the six features, signed so that agreement with the expected direction counts positively, and the highest-scoring factor was selected. Factor sign is arbitrary in latent-variable models, so the selected factor was oriented such that higher values denote the acute host-response state. Associations between factor values and individual features were estimated by linear mixed-effects models with sampling day as a fixed effect and patient as a random intercept.

### Consensus endotype assignments

The consensus transcriptomic subtype (CTS) and consensus immune dysregulation frameworks were applied to sample-level pseudobulk formed by summing raw unique molecular identifier counts across all cells from one patient-day. For the consensus transcriptomic subtype, assignments were made using the *run_subtype_classifier()* function from the ConsensusTranscriptomicSubtype R package^18^, which applies a random-forest classifier to the core classifier genes. For the consensus immune dysregulation framework, the myeloid score was the geometric mean of the myeloid detrimental genes minus the geometric mean of the myeloid protective genes; because the framework defines no lymphoid detrimental gene set, the lymphoid score was the negative geometric mean of the lymphoid protective genes, so that higher values of both scores denote greater dysregulation^19^. Scores were expressed as z-scores relative to the healthy controls of this cohort, and samples were assigned to quadrants at z = 1.65 on both axes, with endotype characterization as balanced, myeloid dysregulation, lymphoid dysregulation and system-wide dysregulation. Endotype stability was assessed among patients contributing more than one sample as the proportion changing class between day 0 and any later acute timepoint, with Wilson score confidence intervals. Transitions out of and into CTS1, the lymphoid dysregulation class and plasma proteomic state 2 were counted separately and compared by exact binomial test.

### Reanalysis of an independent whole-blood cohort

To determine whether the monocyte kinetics observed in this study have a counterpart in granulocytes, we reanalyzed the published whole-blood single-cell sepsis cohort of Kwok *et al.*^21^. CD14^+^ monocytes were reclustered and reannotated to resolve an MS1 state, which the original annotation did not separate; IL1R2^+^ immature neutrophils and other cell states were taken as annotated in that study. Abundances were expressed on the leukocyte denominator for states that include granulocytes and on the mononuclear denominator otherwise. Healthy control, acute sepsis and convalescent groups were compared pairwise by two-sided Wilcoxon test, paired for the nine participants contributing both acute and convalescent samples and unpaired otherwise, with Benjamini–Hochberg adjustment applied across all states tested on a given denominator. The three pairwise comparisons are reported separately rather than pooled, because they are not independent.

### Clinical associations and outcome analyses

Two clinical endpoints, illness severity at presentation and in-hospital mortality, were applied to every immune measure. Illness severity at presentation, measured by the day 0 SOFA score, was chosen because the presentation sample is the one on which enrollment and the earliest treatment decisions would rest. In-hospital mortality was chosen over later endpoints because deaths during the index admission are the most plausibly attributable to the sepsis episode itself. For measures obtained serially, survivors and patients who died in hospital were compared at each timepoint separately, because an association with outcome may itself depend on when the response is measured. Associations of the MS1 fraction, the PBMC SRS score and the latent host response factor with illness severity were estimated as Spearman correlations against the day-matched SOFA score, and pooled across the course by linear mixed-effects models with sampling day as a fixed effect and patient as a random intercept. Comparisons between survivors and patients who died in hospital were made at each timepoint separately by two-sided Wilcoxon rank-sum test; convalescent samples were excluded from these comparisons, because only patients who survived to discharge could return for convalescent sampling, so that group contains no non-survivors to compare against. Within-patient resolution was assessed as the change in each measure between presentation and day 3, correlated against the parallel change in SOFA score.

### Sensitivity of healthy-control-referenced contrasts

Because the healthy control group is small and sampled once, every principal contrast referenced to healthy controls was re-estimated 12 times, omitting one control on each occasion. For each contrast we report the full-model estimate and adjusted P value, the fraction of iterations preserving the sign and the significance of the effect, and the median and 5th to 95th percentile range of the leave-one-out estimates.

### Statistical analysis

Cross-sectional comparisons of the three groups used linear models adjusted for age and sex, with Tukey-adjusted pairwise contrasts. Longitudinal analyses used linear mixed-effects models with patient as a random intercept and age and sex as fixed covariates. Comparisons of two groups on a continuous measure used the two-sided Wilcoxon rank-sum test, comparisons of more than two used the Kruskal–Wallis test, and comparisons of proportions used Fisher’s exact test. Multiple comparisons were otherwise controlled by the Benjamini–Hochberg method. All *P* values are two-sided; *P_adj_* denotes a value adjusted for multiple comparisons, by Tukey’s method for pairwise contrasts among the three groups and by the Benjamini–Hochberg method otherwise. No statistical method was used to predetermine sample size; the cohort comprises all eligible participants enrolled during the study period. No data were excluded other than samples failing quality-control criteria. The study is observational and participants were not randomized.

## DATA AND CODE AVAILABILITY

Raw sequencing data will be deposited under controlled access; processed count matrices and the annotated single-cell object, the plasma proteomic matrix and the sample-level analysis table will be made available with publication. Analysis code will be released as a repository to github.com/pankomah.

## ACKNOWLEDGEMENTS

This work was supported in part by the National Institutes of Health (NIH R01AI153142 to MBG, NH, RPB, PCB, POA, AS, AD, MR), NIH K08 AI185298-01A1 (to POA), NIH KL2 5KL2TR002542-05 (to POA via Harvard Medical School), NIH T32AI007061 (to POA), NIH T32HL155020 (to ALY). The content is solely the responsibility of the authors and does not necessarily represent the official views of the National Institutes of Health.

